# Effectiveness of COVID-19 treatment with nirmatrelvir-ritonavir or molnupiravir among U.S. Veterans: target trial emulation studies with one-month and six-month outcomes

**DOI:** 10.1101/2022.12.05.22283134

**Authors:** Kristina L. Bajema, Kristin Berry, Elani Streja, Nallakkandi Rajeevan, Yuli Li, Lei Yan, Francesca Cunningham, Denise M. Hynes, Mazhgan Rowneki, Amy Bohnert, Edward J. Boyko, Theodore J. Iwashyna, Matthew L. Maciejewski, Thomas F. Osborne, Elizabeth M. Viglianti, Mihaela Aslan, Grant D. Huang, George N. Ioannou

## Abstract

**Background:** Information about the effectiveness of oral antivirals in preventing short- and long-term COVID-19-related outcomes during the Omicron surge is limited. We sought to determine the effectiveness of nirmatrelvir-ritonavir and molnupiravir for the outpatient treatment of COVID-19.

**Methods:** We conducted three retrospective target trial emulation studies comparing matched patient cohorts who received nirmatrelvir-ritonavir versus no treatment, molnupiravir versus no treatment, and nirmatrelvir-ritonavir versus molnupiravir in the Veterans Health Administration (VHA). Participants were Veterans in VHA care at risk for severe COVID-19 who tested positive for SARS-CoV-2 in the outpatient setting during January and February 2022. Primary outcomes included all-cause 30-day hospitalization or death and 31-180-day incidence of acute or long-term care admission, death, or post-COVID-19 conditions. For 30-day outcomes, we calculated unadjusted risk rates, risk differences, and risk ratios. For 31-180-day outcomes, we used unadjusted time-to-event analyses.

**Results:** Participants were 90% male with median age 67 years and 26% unvaccinated. Compared to matched untreated controls, nirmatrelvir-ritonavir-treated participants (N=1,587) had a lower 30-day risk of hospitalization (27.10/1000 versus 41.06/1000, risk difference [RD] - 13.97, 95% CI -23.85 to -4.09) and death (3.15/1000 versus 14.86/1000, RD -11.71, 95% CI - 16.07 to -7.35). Among persons who were alive at day 31, further significant reductions in 31-180-day incidence of hospitalization (sub-hazard ratio 1.07, 95% CI 0.83 to 1.37) or death (hazard ratio 0.61, 95% CI 0.35 to 1.08) were not observed. Molnupiravir-treated participants aged ≥65 years (n=543) had a lower combined 30-day risk of hospitalization or death (55.25/1000 versus 82.35/1000, RD -27.10, 95% CI -50.63 to -3.58). A statistically significant difference in 30-day or 31-180-day risk of hospitalization or death was not observed between matched nirmatrelvir- or molnupiravir-treated participants. Incidence of most post-COVID conditions was similar across comparison groups.

**Conclusions:** Nirmatrelvir-ritonavir was highly effective in preventing 30-day hospitalization and death. Short-term benefit from molnupiravir was observed in older groups. Significant reductions in adverse outcomes from 31-180 days were not observed with either antiviral.

## INTRODUCTION

Two pharmacotherapies, nirmatrelvir packaged with the boosting agent ritonavir (nirmatrelvir-ritonavir) as well as molnupiravir, received emergency use authorization by the U.S. Food and Drug Administration (FDA) in December 2021 for the treatment of non-hospitalized persons with symptomatic COVID-19 at high risk for progression to severe COVID-19. Early randomized controlled trials (RCTs) demonstrated a reduction in COVID-19-related hospitalization or death with nirmatrelvir-ritonavir or molnupiravir compared to placebo (1, 2).

Effectiveness studies of nirmatrelvir-ritonavir and molnupiravir are needed because clinical trials were conducted among unvaccinated participants before the emergence of the Omicron (B.1.1.529) and subsequent sublineages. The RCTs did not directly compare efficacy of antiviral agents, nor did they evaluate outcomes beyond 29 days following symptomatic infection. The effect of these antivirals on post-acute complications of SARS-CoV-2 is also unknown. Early observational studies of nirmatrelvir-ritonavir (3-6) and molnupiravir (7, 8) have demonstrated reduced risk of short-term hospitalization and death. Non-interventional studies adhering to target trial emulation principles (9) are needed to carefully evaluate whether these antivirals are effective against the now predominant Omicron variants especially in older, racially, and ethnically diverse populations with a high prevalence of underlying conditions.

The Veterans Health Administration (VHA), run by the U.S. Department of Veterans Affairs (VA), is the largest integrated healthcare system in the U.S. providing care to more than 9 million Veterans with a majority of older age and high burden of underlying medical conditions, has provided an opportunity for multiple target trial emulation studies of the comparative effectiveness of COVID-19 pharmacotherapies and vaccines (10-13). We used target trial emulation principles (9) to emulate three trials including nirmatrelvir-ritonavir versus no treatment, molnupiravir versus no treatment, and nirmatrelvir-ritonavir versus molnupiravir during the early Omicron era (B.1.1.529 and BA1.1). We evaluated acute 30-day hospitalization and death outcomes as well as 6-month incidence of acute or long-term care admission, death, and post-COVID conditions among non-hospitalized adult Veterans infected with SARS-CoV-2 at high risk for progression to severe COVID-19.

## METHODS

### Specification and emulation of target trials: overall study design

We designed this retrospective cohort study to emulate three target randomized controlled trials of COVID-19 antiviral agents among symptomatic, non-hospitalized adult Veterans enrolled in VHA with a first positive SARS-CoV-2 test from January 1 through February 28, 2022 who were at high risk for progression to severe COVID-19: nirmatrelvir-ritonavir versus no SARS-CoV-2 antiviral or monoclonal antibody treatment (trial 1); molnupiravir versus no treatment (trial 2); and nirmatrelvir-ritonavir versus molnupiravir (trial 3). The follow-up period extended through August 31, 2022, to allow ascertainment of short-term, 30-day outcomes and long-term, 6-month outcomes following treatment. Target trial emulation applies design principles from randomized trials to the analysis of observational data, thereby explicitly tying the design and analysis to the hypothetical trial it is emulating (14). **Supplemental Table 1** compares the critical study design features of the specified and emulated target trials (9). We used a matched cohort design to emulate the balance achieved through randomization. Untreated persons were assigned an index date that was the same number of days after the date of the first positive SARS-CoV-2 test (test-positive date) as the treatment date of the matched treated patients (**Figure 1**).

**Figure 1.**
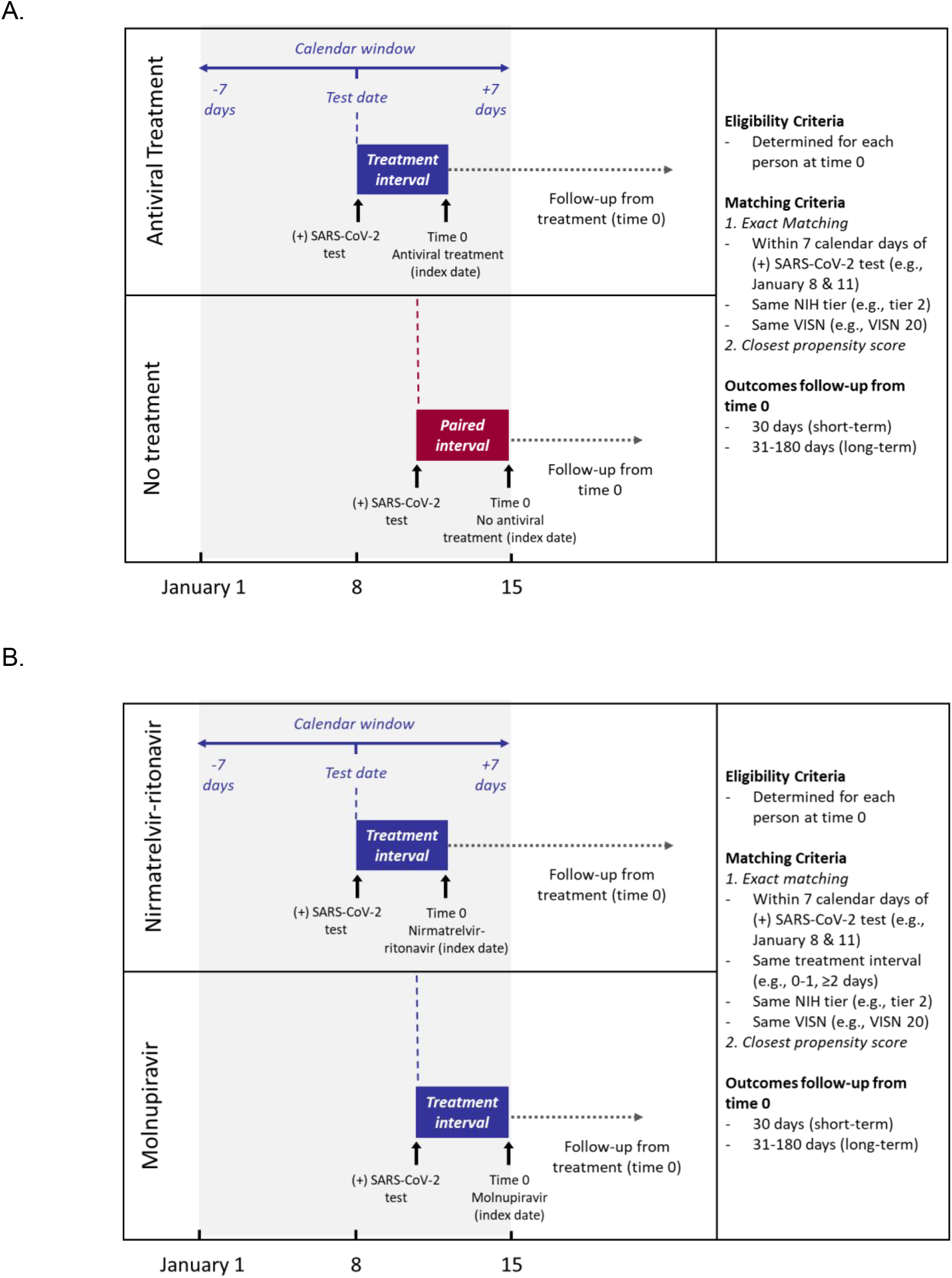
Matching strategy for A. target trials 1 and 2 (nirmatrelvir versus no treatment and molnupiravir versus no treatment) and B. target trial 3 (nirmatrelvir versus molnupiravir).

Eligibility criteria were ascertained as of this index date, and follow-up for each matched set began as of this index date (time zero) and continued until occurrence of an outcome event or the end of the 6-month follow-up period. The study was approved by the VA Central Institutional Review Board and followed the Strengthening the Reporting of Observational Studies in Epidemiology (STROBE) reporting guideline.

### Data sources

We used VHA’s COVID-19 Shared Data Resource (CSDR), supported by the VA Informatics and Computing Infrastructure (VINCI), which integrates multiple data sources to provide patient-level COVID-19-related information on VHA enrollees. CSDR includes information on laboratory-confirmed, positive, SARS-CoV-2 tests (either by nucleic acid amplification or antigen testing) within the VHA system as well as SARS-CoV-2 tests performed outside VHA and documented in VHA clinical records. Positive tests are identified by the VA National Surveillance Tool and provisioned to the CSDR to support national VA research and operational needs.

These data were supplemented with detailed claims data from the VA Community Care program, which coordinates and reimburses VA purchased care provided in the community and from the Centers for Medicare and Medicaid Services (CMS), provisioned by the VA Information Resource Center (VIReC). VA Community care and CMS-Medicare data were used to capture additional COVID-19 antiviral or monoclonal antibody treatments (i.e., nirmatrelvir-ritonavir, molnupiravir, sotrovimab, remdesivir), COVID-19 vaccinations, hospitalizations, and post-COVID conditions.

### Eligibility criteria and study population

We identified all VHA enrollees aged 18 years and older with a first positive SARS-CoV-2 test in CSDR from January 1-February 28, 2022 (**Figure 2**). We limited the study population to VHA enrollees with a VHA primary care encounter in the 18 months preceding the test-positive date who were alive and not hospitalized on or within 7 days before the test-positive date. Treated participants who died or were hospitalized on or before their antiviral treatment date were also excluded; identical exclusions for untreated participants relative to their assigned index date were later applied during the matching process. We excluded persons who received nirmatrelvir-ritonavir or molnupiravir before or more than 10 days after the test-positive date, persons receiving a different outpatient COVID-19 treatment within 7 days prior to the antiviral treatment date (**Supplemental Methods**), and persons who did not have at least one risk factor for progression to severe COVID-19 (**Supplemental Table 2**,**3**) (15). For comparisons involving nirmatrelvir-ritonavir, we excluded persons with advanced renal or hepatic disease as well as persons with absolute drug contraindications (**Supplemental Methods, Supplemental Table 4**) (16). For comparisons involving molnupiravir, we excluded pregnant persons. Persons were eligible as untreated comparators if they did not receive any outpatient COVID-19 pharmacotherapies on or before their assigned index date.

**Figure 2.**
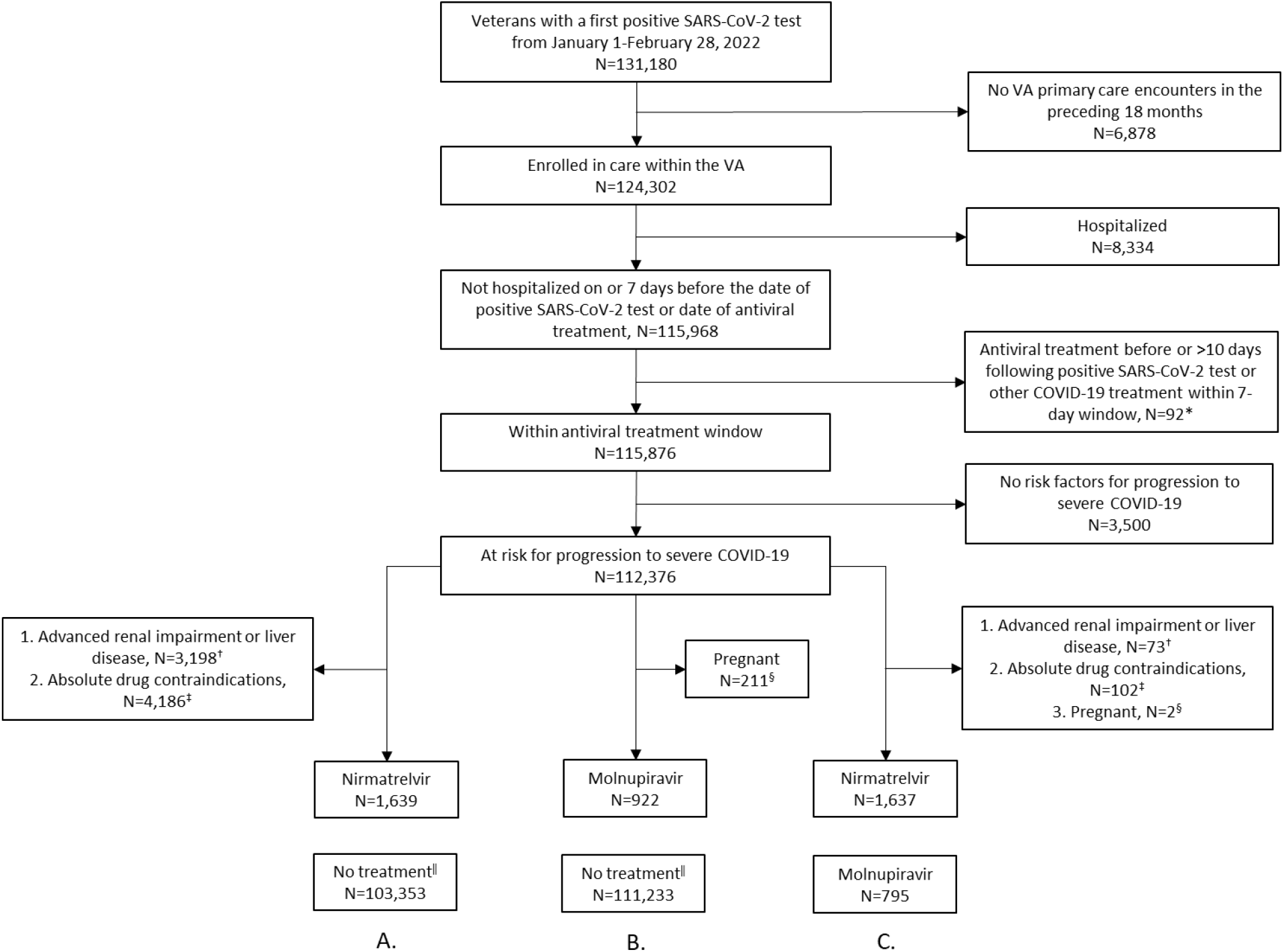
Criteria for the identification of eligible study participants for the emulation of three target trials comparing the effectiveness of A. nirmatrelvir-ritonavir versus no treatment, B. molnupiravir versus no treatment, and C. nirmatrelvir-ritonavir versus molnupiravir.

^*^Excludes persons receiving COVID-19 antiviral agents (nirmatrelvir-ritonavir or molnupiravir) outside of an expected treatment window, allowing for small discrepancies in test-positive and treatment dates. Also excludes persons who received other outpatient COVID-19 treatments (nirmatrelvir-ritonavir, molnupiravir, sotrovimab, remdesivir) on or prior to the antiviral treatment date.

^†^See Supplemental Methods.

^‡^See Supplemental Table 4.

^§^Documented within 1 week prior to positive SARS-CoV-2 test date.

^‖^Match eligible numbers presented here include persons who received nirmatrelvir-ritonavir, molnupiravir, sotrovimab, or remdesivir between January-February 2022. See Supplemental Table 9 for additional exclusions for any treatments received on or prior to the matched index date.

### Cohort matching

Two matching steps were used to achieve balance of covariates between comparator groups and reduce confounding.

#### Exact-matching

We first exact-matched each eligible participant who received nirmatrelvir-ritonavir or molnupiravir to all eligible participants who were untreated as of their assigned index date using three factors: NIH tier of prioritization for anti-SARS-CoV-2 therapies (**Supplemental Table 5**); VA Integrated Service Network (VISN), the 19 geographical administrative regions of the VA; and calendar time, centered within 7 days of the test-positive date of the matched comparator (**Figure 1**). For the comparison of nirmatrelvir-ritonavir versus molnupiravir, additional exact-matching based on test-date to treatment-date interval (0-1 days versus 2-10 days) was used.

#### Propensity-score matching

We performed an additional propensity score matching step with replacement in a 1:k variable ratio, where k varied based on the number of propensity score ties. We included all ties to avoid imbalance due to random pruning. We included in the propensity score logistic regression model predicting treatment the demographic, geographic, healthcare utilization, and clinical factors selected a priori based on their association with both the treatment exposure and outcomes (**Supplemental Tables 6**,**7**). Up to four untreated participants with the closest propensity scores within 0.2 standard deviations of the mean (sdm) were matched to each treated participant. In accordance with an intent-to-treat approach to analysis, assigned untreated participants who later received treatment after the index date were not censored. This approach was also used for participants assigned to nirmatrelvir-ritonavir or molnupiravir groups who may have later received a different pharmacotherapy. Each molnupiravir-treated participant was matched with replacement to the single nirmatrelvir-ritonavir-treated participant with the closest propensity score within 0.4 sdm. Nirmatrelvir-ritonavir-treated participants could serve as matched comparators to more than one molnupiravir-treated participant.

### Primary endpoints

#### Short-term outcomes

The primary short-term outcomes of interest were any hospitalization or all-cause death through day 30 following the index date (time zero). We also evaluated as secondary outcomes ICU admission and mechanical ventilation occurring during hospitalizations through day 30.

#### Long-term outcomes

We determined the 6-month incidence of any acute or long-term care admission (including hospitals and skilled nursing facilities) or all-cause death, measured from 31-180 days among matched groups alive at day 31. We also assessed the incidence of 34 potential post-COVID conditions described in the literature (17-20) (**Supplemental Table 8**) from day 31-180 following the index date. For each condition, analysis was limited to matched groups where all persons were alive at day 30 and did not have the condition of interest documented within 1 year prior to the index date.

### Statistical analyses

Patient characteristics were compared between arms in each of the three trial emulations. For 30-day outcomes of hospitalization or death, we calculated unadjusted risk rates, risk differences, risk ratios (and 95% CIs) and plotted Kaplan-Meier curves. For incidence of long-term outcomes extending from 31-180 days, we used unadjusted time-to-event analyses treating death as a competing risk. Subgroup analyses were considered by age (less than 65 years versus 65 and older), vaccination status (unvaccinated versus any primary or booster vaccination) and presence or absence of COVID-19-related symptoms within 30 days prior to the test-date. Missing or unknown values in Care Assessment Need (CAN) score and race/ethnicity were uncommon and treated as a separate unknown category (21, 22).

All analyses were importance-weighted to account for variable-ratio matching (23). A robust sandwich-type variance estimator was used to account for clustering within the matched group due to ties in the propensity score, clustering within subjects due to matching with replacement, and clustering in the cross-classification of the matched and within subject clusters (24). We verified that the proportional hazards assumption was met using log-log plots and Schoenfeld residuals. A p-value <0.05 was considered statistically significant. Analyses were conducted using STATA (StataCorp).

## RESULTS

### Patient population

A total of 112,380 Veterans with a first positive SARS-CoV-2 test during January and February 2022 were identified for inclusion in our study, of whom 1,639 of 103,353 (1.6%) match-eligible persons received nirmatrelvir-ritonavir in trial 1 and 922 of 111,233 (0.8%) match-eligible persons received molnupiravir in trial 2 (**Figure 2**). In trial 3, 1,637 match-eligible nirmatrelvir-ritonavir recipients and 795 match-eligible molnupiravir recipients were identified. Baseline characteristics were well balanced between the matched comparator groups of each of the three emulated trials with sdms all below 0.10 (**Table 1, Supplemental Figures 1-3**). Matching with replacement allowed matching of 1587 (96.8%) of the eligible nirmatrelvir-ritonavir-treated participants in trial 1, 897 (97.1%) of the eligible molnupiravir-treated participants in trial 2, and 769 (96.7%) of the molnupiravir-treated participants in trial 3 who were matched to 534 unique nirmatrelvir-ritonavir-treated participants (**Supplemental Table 11**).

**Table 1.** Baseline demographic and clinical characteristics of Veterans who tested positive for SARS-CoV-2 in the VA healthcare system from January 1, 2022 to February 28, 2022 who fulfilled eligibility criteria and were matched as participants in three emulated target trials of COVID-19 antiviral effectiveness

| Characteristic | Trial 1 |  | Trial 2 |  | Trial 3 |  |
| --- | --- | --- | --- | --- | --- | --- |
|  | Nirmatrelvir-ritonavir | No treatment* | Molnupiravir | No treatment* | Molnupiravir | Nirmatrelvir-ritonavir |
| **Total** | 1,587 | 1,587 | 897 | 897 | 769 | 769 |
| **Age, years, median [IQR]** | 65.0<br>(54.0,74.0) | 66.0<br>(54.0,74.0) | 68.0<br>(57.0,74.0) | 68.0<br>(57.0,74.0) | 68.0<br>(57.0,74.0) | 67.0<br>(57.0,74.0) |
| **Age group, N (%)** |  |  |  |  |  |  |
| 18-49 | 279(17.6) | 290(18.3) | 110(12.3) | 121(13.5) | 101(13.1) | 103(13.4) |
| 50-64 | 464(29.2) | 443(27.9) | 244(27.2) | 238(26.5) | 215(28.0) | 219(28.5) |
| 65-74 | 522(32.9) | 512(32.3) | 322(35.9) | 318(35.4) | 271(35.2) | 272(35.4) |
| ≥75 | 322(20.3) | 341(21.5) | 221(24.6) | 220(24.6) | 182(23.7) | 175(22.8) |
| **Male sex, N (%)** | 1,412(89.0) | 1,416(89.3) | 815(90.9) | 818(91.2) | 693(90.1) | 690(89.7) |
| **Race/Ethnicity, N (%)** |  |  |  |  |  |  |
| Hispanic | 142(8.9) | 123(7.7) | 54(6.0) | 46(5.2) | 46(6.0) | 51(6.6) |
| White | 1,111(70.0) | 1,149(72.4) | 659(73.5) | 675(75.2) | 573(74.5) | 551(71.7) |
| Black | 231(14.6) | 222(14.0) | 129(14.4) | 112(12.5) | 99(12.9) | 115(15.0) |
| Other | 38(2.4) | 35(2.2) | 19(2.1) | 21(2.4) | 17(2.2) | 20(2.6) |
| Unknown | 65(4.1) | 59(3.7) | 36(4.0) | 43(4.8) | 34(4.4) | 32(4.2) |
| **Rurality, N (%)†** |  |  |  |  |  |  |
| Rural | 293(18.5) | 316(19.9) | 196(21.9) | 210(23.4) | 164(21.3) | 145(18.9) |
| Urban | 1,294(81.5) | 1,271(80.1) | 701(78.1) | 687(76.6) | 605(78.7) | 624(81.1) |
| **Region, N (%)‡** |  |  |  |  |  |  |
| West | 337(21.2) | 337(21.2) | 223(24.9) | 223(24.9) | 194(25.2) | 194(25.2) |
| Midwest | 397(25.0) | 397(25.0) | 202(22.5) | 202(22.5) | 162(21.1) | 162(21.1) |
| Northeast | 308(19.4) | 308(19.4) | 90(10.0) | 90(10.0) | 77(10.0) | 77(10.0) |
| South | 545(34.3) | 545(34.3) | 382(42.6) | 382(42.6) | 336(43.7) | 336(43.7) |
| **Area Deprivation Index, median [IQR]** | 59.3<br>(41.0,76.8) | 60.3<br>(42.3,76.7) | 61.2<br>(43.7,77.7) | 62.5<br>(45.1,76.7) | 62.3<br>(45.0,78.2) | 59.4<br>(41.5,75.7) |
| **Week of positive test, N (%)** |  |  |  |  |  |  |
| January 1-14 | 188 (11.9) | 169 (10.7) | 137 (15.3) | 118 (13.2) | 113 (14.7) | 96 (12.5) |
| January 15-28 | 730 (46.0) | 765 (48.2) | 452 (50.4) | 482 (53.7) | 399 (51.9) | 441 (57.3) |
| January 29-February 11 | 456 (28.8) | 452 (28.5) | 226 (25.2) | 221 (24.7) | 185 (24.0) | 162 (21.1) |
| February 12-28 | 213 (13.4) | 202 (12.7) | 82 (9.2) | 85 (8.4) | 72 (9.4) | 70.0 (9.1) |
| **≥1 symptom, N (%)§** |  |  |  |  |  |  |
| No | 461(29.0) | 470(29.6) | 219(24.4) | 222(24.7) | 190(24.7) | 151(19.6) |
| Yes | 1,126(71.0) | 1,117(70.4) | 678(75.6) | 675(75.3) | 579(75.3) | 618(80.4) |
| **Vaccination status and time since last dose, N (%)** |  |  |  |  |  |  |
| No doses | 461(29.0) | 479(30.2) | 201(22.4) | 211(23.6) | 182(23.7) | 184(23.9) |
| Partial | 76 (4.7) | 73 (4.6) | 37 (4.1) | 34 (3.8) | 29 (3.8) | 33 (4.3) |
| Primary/>4 months | 489(30.8) | 476(30.0) | 304(33.9) | 301(33.6) | 264(34.3) | 274(35.6) |
| Primary/0-4 months | 72(4.5) | 61(3.9) | 39(4.3) | 31(3.5) | 33(4.3) | 28(3.6) |
| Booster/>4 months | 98(6.2) | 87(5.5) | 60(6.7) | 50(5.6) | 51(6.6) | 63(8.2) |
| Booster/0-4 months | 391(24.6) | 411(25.9) | 256(28.5) | 270(30.1) | 209(27.2) | 187(24.3) |
| **NIH Tier, N (%)** |  |  |  |  |  |  |
| 1 | 327(20.6) | 327(20.6) | 156(17.4) | 156(17.4) | 135(17.6) | 135(17.6) |
| 2 | 276(17.4) | 276(17.4) | 115(12.8) | 115(12.8) | 106(13.8) | 106(13.8) |
| 3 | 589(37.1) | 589(37.1) | 404(45.0) | 404(45.0) | 334(43.4) | 334(43.4) |
| 4 | 395(24.9) | 395(24.9) | 222(24.7) | 222(24.7) | 194(25.2) | 194(25.2) |
| **Smoking, N (%)** |  |  |  |  |  |  |
| Never | 625(39.4) | 607(38.3) | 324(36.1) | 309(34.5) | 278(36.2) | 270(35.1) |
| Former | 672(42.3) | 653(41.2) | 397(44.3) | 416(46.3) | 340(44.2) | 355(46.2) |
| Current | 233(14.7) | 266(16.8) | 152(16.9) | 147(16.4) | 130(16.9) | 127(16.5) |
| Unknown | 57(3.6) | 61(3.8) | 24(2.7) | 25(2.8) | 21(2.7) | 17(2.2) |
| **Alcohol dependence, N (%)** | 281(17.7) | 294(18.5) | 157(17.5) | 158(17.6) | 133(17.3) | 157(20.4) |
| **Substance dependence, N (%)** | 56(3.5) | 52(3.3) | 37(4.1) | 34(3.8) | 30(3.9) | 28(3.6) |
| **Number of underlying conditions** |  |  |  |  |  |  |
| **Median [IQR]** | 4.0 (2.0,6.0) | 4.0 (2.0,6.0) | 4.0 (3.0,14.0) | 4.0<br>(3.0,14.0) | 4.0 (3.0,13.0) | 4.0 (3.0,13.0) |
| **N (%)** |  |  |  |  |  |  |
| 0-1 | 152(9.6) | 173(10.9) | 50(5.6) | 68(7.6) | 48(6.2) | 44(5.7) |
| 2-3 | 618(38.9) | 619(39.0) | 281(31.3) | 294(32.8) | 255(33.2) | 291(37.8) |
| 4-5 | 356(22.4) | 341(21.5) | 212(23.6) | 196(21.8) | 194(25.2) | 172(22.4) |
| 5+ | 461(29.0) | 454(28.6) | 354(39.5) | 339(37.8) | 272(35.4) | 262(34.1) |
| **CAN Score for mortality w/in** |  |  |  |  |  |  |
| **1yr at test date (%)<sup>†</sup>** |  |  |  |  |  |  |
| 0-30 | 497(31.3) | 500(31.5) | 216(24.1) | 218(24.3) | 193(25.1) | 187(24.3) |
| 31-55 | 438(27.6) | 438(27.6) | 230(25.6) | 239(26.6) | 207(26.9) | 234(30.4) |
| 56-75 | 302(19.0) | 313(19.7) | 195(21.7) | 197(22.0) | 179(23.3) | 169(22.0) |
| 76-90 | 235(14.8) | 234(14.7) | 170(19.0) | 162(18.0) | 136(17.7) | 126(16.4) |
| 95-96 | 45(2.8) | 36(2.3) | 26(2.9) | 25(2.8) | 17(2.2) | 18(2.3) |
| 97-98 | 36(2.3) | 32(2.0) | 37(4.1) | 34(3.8) | 21(2.7) | 20(2.6) |
| 99 | 22(1.4) | 19(1.2) | 21(2.3) | 18(2.0) | 15(2.0) | 14(1.8) |
| **Underlying condition, N (%)** |  |  |  |  |  |  |
| Obesity (body mass index $\geq 30$ kg/m<sup>2</sup>) | 818(51.5) | 817(51.5) | 476(53.1) | 484(53.9) | 405(52.7) | 423(55.0) |
| Chronic kidney disease | 187(11.8) | 189(11.9) | 185(20.6) | 171(19.0) | 111(14.4) | 105(13.7) |
| Diabetes | 594(37.4) | 576(36.3) | 375(41.8) | 362(40.3) | 301(39.1) | 301(39.1) |
| Immuno-suppressive medications or cancer therapies<sup>‡</sup> | 143(9.0) | 133(8.4) | 65(7.2) | 60(6.7) | 57(7.4) | 50(6.5) |
| Cancer | 406(25.6) | 335(21.1) | 226(25.2) | 214(23.8) | 181(23.5) | 206(26.8) |
| Cardiovascular disease | 555(35.0) | 556(35.0) | 430(47.9) | 399(44.5) | 334(43.4) | 304(39.5) |
| Chronic lung disease | 547(34.5) | 534(33.7) | 365(40.7) | 346(38.6) | 308(40.1) | 292(38.0) |
| Dementia | 59(3.7) | 55(3.5) | 44(4.9) | 41(4.6) | 34(4.4) | 28(3.6) |
| Cerebrovascular disease | 78(4.9) | 90(5.7) | 82(9.1) | 68(7.6) | 62(8.1) | 45(5.9) |
| Chronic liver disease | -- | -- | 5(0.6) | 6(0.7) | -- | -- |
| Mental Health conditions<sup>***</sup> | 693(43.7) | 742(46.8) | 441(49.2) | 402(44.8) | 375(48.8) | 336(43.7) |
| **Number of healthcare encounters in prior 12 months, mean±SD** | 26.2±24.6 | 25.6±22.8 | 30.4±28.8 | 29.3±27.1 | 27.4±23.1 | 26.8±23.3 |
| **Number of healthcare encounters in prior 12 months, median (IQR)** | 19.0<br>(11.0,34.0) | 19.0<br>(11.0,34.0) | 23.0<br>(13.0,37.0) | 22.0<br>(12.0,38.0) | 22.0<br>(12.0,35.0) | 21.0<br>(13.0,35.0) |
| **Number of healthcare encounters in prior 12 months, N (%)** |  |  |  |  |  |  |
| 0-8 | 286(18.0) | 294(18.5) | 124(13.8) | 136(15.1) | 117(15.2) | 112(14.6) |
| 9-15 | 318(20.0) | 327(20.6) | 163(18.2) | 167(18.6) | 147(19.1) | 157(20.4) |
| 16-30 | 517(32.6) | 509(32.1) | 300(33.4) | 290(32.3) | 264(34.3) | 266(34.6) |
| 30+ | 466(29.4) | 456(28.8) | 310(34.6) | 304(33.9) | 241(31.3) | 234(30.4) |
| **Days from test to treatment, N (%)** |  |  |  |  |  |  |
| 0-1 | 1304(82.2) | - | 746(83.2) | - | 652(84.8) | 652(84.8) |
| 2-10 | 283(17.8) | - | 151(16.8) | - | 117(15.2) | 117(15.2) |

CAN = Care Assessment Need; NIH = National Institutes of Health
\*Baseline characteristics represent equally weighted matched controls (up to 4 untreated persons for trials 1 and 2)
†Based on rural-urban commuting area (RUCA) codes
‡Regions are based on Veterans Integrated Service Networks (VISNs). West includes VISNs 19-22; Midwest 10,12,15,23; Northeast 1,2,4,5; South 6-9, 16-17
§Any of 15 pre-specified COVID-19-related symptoms present on the day of positive SARS-CoV-2 test or within the preceding 30 days
||See Supplemental Appendix
¶CAN data missing for 35 persons
\*\*Includes major depressive disorder, bipolar disorder, post-traumatic stress disorder, schizophrenia

Across all matched groups in the three trials, participants were mostly male (range 89.0-91.2%) with advanced median age (range 65-68 years), 5.2-8.9% Hispanic ethnicity, 70.0-75.2% White race, 12.5-15.0% Black race, and a median of 4 medical conditions associated with severe COVID-19 risk (15) led by obesity (range 51.5-55.0%), mental health conditions (43.7-49.2%), and cardiovascular disease (range 35.0-47.9%) (**Table 1**). Overall, 22.4-30.2% of participants were not vaccinated for COVID-19.

### Short-term outcomes

#### Nirmatrelvir-ritonavir versus no treatment

The 30-day rate of hospitalization or death was significantly lower in the nirmatrelvir-ritonavir arm (28.36 events per 1000 persons) than the no treatment arm (53.40 events per 1000 persons), a rate difference (RD) of -25.05 events per 1000 persons (95% CI -35.47 to -14.62) and risk ratio (RR) 0.53 (95% CI 0.39 to 0.72, **Figure 3, Table 2**), which was driven by persons aged 65 years and older (RD -42.29 events per 1000 persons, 95% CI -58.61 to -25.97; RR 0.46, 95% CI 0.31 to 0.66, **Supplemental Tables 12, 13**). Overall, there was a reduction in both death (RD -11.71 events per 1000 persons, 95% CI -16.07 to -7.35; RR 0.21, 95% CI 0.09 to 0.52) and hospitalization (RD -13.97 events per 1000 persons, 95% CI -23.85 to -4.09; RR 0.66, 95% CI 0.48 to 0.91).

**Figure 3.**
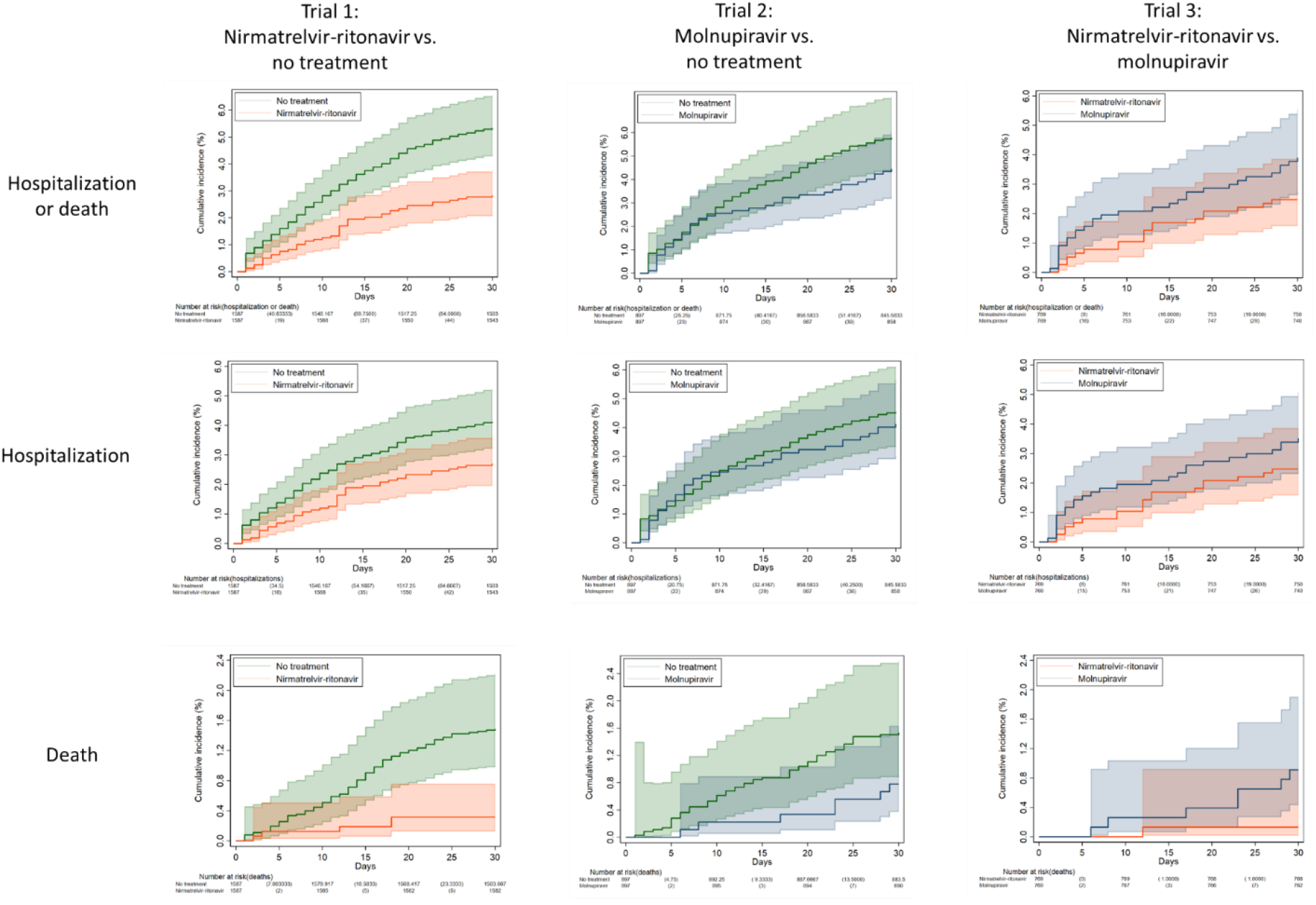
Kaplan-Meier curves comparing Veterans treated with nirmatrelvir-ritonavir versus their matched untreated counterparts (trial 1), molnupiravir versus their matched untreated counterparts (trial 2), and nirmatrelvir-ritonavir versus molnupiravir (trial 3) showing cumulative incidence (%) with 95% confidence intervals of any hospitalization or all-cause death, any hospitalization, and all-cause death through day 30 after the index date.

**Table 2.** Comparison of matched groups in three emulated target trials of COVID-19 pharmacotherapy among Veterans who tested positive for SARS-CoV-2 from January 1, 2022 to February 28, 2022 with respect to 30-day outcomes of death and/or hospitalization, ICU admission and mechanical ventilation after index date

| 30-day Outcome | No. of Events |  | 30-day risk, events per 1000 persons (95% CI*) |  | Risk Difference (95% CI*) | Risk Ratio (95% CI*) |
| --- | --- | --- | --- | --- | --- | --- |
| Trial #1 | Nirmatrelvir-ritonavir | No treatment | Nirmatrelvir-ritonavir | No treatment |  |  |
| Hospitalization or Death | 45 | 84.8 | 28.36<br>(21.23-37.78) | 53.40<br>(47.31-60.23) | -25.05<br>(-35.47--14.62) | 0.53<br>(0.39-0.72) |
| Hospitalization | 43 | 65.2 | 27.10<br>(20.15-36.35) | 41.06<br>(35.61-47.30) | -13.97<br>(-23.85--4.09) | 0.66<br>( 0.48- 0.91) |
| Death | 5 | 23.6 | 3.15<br>( 1.31- 7.55) | 14.86<br>(11.93-18.50) | -11.71<br>(-16.07--7.35) | 0.21<br>( 0.09- 0.52) |
| ICU Admission | † | 22.8 | 6.30<br>( 3.39-11.68) | 14.39<br>(11.11-18.61) | -8.09<br>(-13.56--2.61) | 0.44<br>( 0.22- 0.86) |
| Mechanical Ventilation | † | 5.8 | 1.89<br>( 0.61- 5.85) | 3.68<br>( 2.29- 5.89) | -1.79<br>(-4.54- 0.97) | 0.51<br>( 0.15- 1.75) |
| **Trial #2** | **Molnupiravir** | **No treatment** | **Molnupiravir** | **No treatment** |  |  |
| Hospitalization or Death | 40 | 51.9 | 44.59<br>(32.86-60.26) | 57.88<br>(49.88-67.07) | -13.29<br>(-29.44- 2.87) | 0.77<br>( 0.55- 1.08) |
| Hospitalization | 37 | 40.5 | 41.25<br>(30.01-56.45) | 45.15<br>(38.07-53.48) | -3.90<br>(-19.07-11.26) | 0.91<br>( 0.64- 1.31) |
| Death | 7 | 13.8 | 7.80<br>( 3.72-16.30) | 15.33<br>(11.53-20.35) | -7.53<br>(-14.89--0.16) | 0.51<br>( 0.23- 1.13) |
| ICU Admission | † | 10.1 | 11.15<br>( 6.00-20.62) | 11.24<br>( 7.81-16.16) | -0.09<br>(-7.99- 7.81) | 0.99<br>( 0.49- 2.01) |
| Mechanical Ventilation | † | 3.6 | 3.34<br>( 1.08-10.34) | 3.99<br>( 2.36- 6.75) | -0.65<br>(-4.98- 3.68) | 0.84<br>( 0.24- 2.92) |
| **Trial #3** | **Nirmatrelvir-ritonavir** | **Molnupiravir** | **Nirmatrelvir-ritonavir** | **Molnupiravir** |  |  |
| Hospitalization or Death | 19 | 30 | 24.71<br>(14.45-41.95) | 39.01<br>(27.39-55.29) | -14.30<br>(-33.20- 4.59) | 0.63<br>( 0.34- 1.19) |
| Hospitalization | 19 | 27 | 24.71<br>(14.45-41.95) | 35.11<br>(24.17-50.75) | -10.40<br>(-28.79- 7.98) | 0.70<br>( 0.37- 1.34) |
| Death | 1 | 7 | 1.30<br>( 0.18- 9.24) | 9.10<br>( 4.34-19.00) | -7.80<br>(-15.00--0.61) | 0.14<br>( 0.02- 1.16) |
| ICU Admission | † | 5 | 5.20<br>( 1.56-17.20) | 6.50<br>( 2.70-15.55) | -1.30<br>(-9.76- 7.16) | 0.80<br>( 0.18- 3.54) |
| Mechanical Ventilation | † | 3 | 1.30<br>( 0.18- 9.24) | 3.90<br>( 1.26-12.05) | -2.60<br>(-7.70- 2.50) | 0.33<br>( 0.03- 3.21) |

ICU = intensive care unit
\*CI accounting for clustering: patient ID for 30-day risk, and patient ID and match ID for risk difference and ratio
†Data from Centers for Medicare & Medicaid Services (CMS) were combined with other data sources and masked in accordance with CMS cell size suppression policy (https://resdac.org/articles/cms-cell-size-suppression-policy)

#### Molnupiravir versus no treatment

The 30-day rate of hospitalization or death was similar between molnupiravir-treated (44.59 events per 1000 persons) and untreated groups (57.88 events per 1000 persons) overall, with a RD of -13.29 events per 1000 persons (95% CI -29.44 to 2.87) and RR 0.77 (95% CI 0.55 to 1.08, **Figure 3, Table 2**). However, among persons aged 65 years and older, receipt of molnupiravir was associated with a lower 30-day risk of hospitalization or death (RD -27.10 events per 1000 persons, 95% CI -50.63 to -3.58; RR 0.67, 95% CI 0.46 to 0.99, **Supplemental Tables 12, 13**). There was a significant reduction in absolute risk of death (RD -7.53 events per 1000 persons, 95% CI -14.89 to -0.16) although the relative risk was not significant (RR 0.51, 95% CI 0.23 to 1.13).

#### Molnupiravir versus nirmatrelvir-ritonavir

The 30-day rate of hospitalization or death was 39.01 events per 1000 persons in the molnupiravir arm compared with 24.71 events per 1000 persons in the nirmatrelvir-ritonavir arm, an insignificant rate difference of -14.30 events per 1000 persons (95% CI -33.20 to 4.59, **Figure 3, Table 2**). Compared with molnupiravir-treated participants, there was a significant reduction in absolute risk of death among nirmatrelvir-ritonavir-treated participants (RD -7.89 events per 1000 persons, 95% CI -15.00 to -0.61) although the relative risk was not significant (RR 0.14, 95% CI 0.02 to 1.16). There were no significant differences in hospitalization.

### Long-term outcomes

There was no significant difference in 31-180-day incidence of acute or long-term care admission or death between nirmatrelvir-ritonavir and no treatment groups (**Figure 4, Table 3**). Incidence of acute or long-term care admission was 123.0 events per 1000 persons in the molnupiravir arm compared with 82.5 events per 1000 persons in the no treatment arm (subhazard ratio [SHR] 1.48, 95% CI 1.11 to 1.99). There was no significant difference in mortality (SHR 1.08, 95% CI 0.62 to 1.89) between these groups. Compared to the no treatment arm, the nirmatrelvir-ritonavir arm had a lower incidence of renal conditions (44.2 versus 65.1 events per 100 persons, SHR 0.68, 95% CI 0.48 to 0.95). There were otherwise no significant differences between groups regarding the incidence of post-COVID conditions.

**Figure 4.**
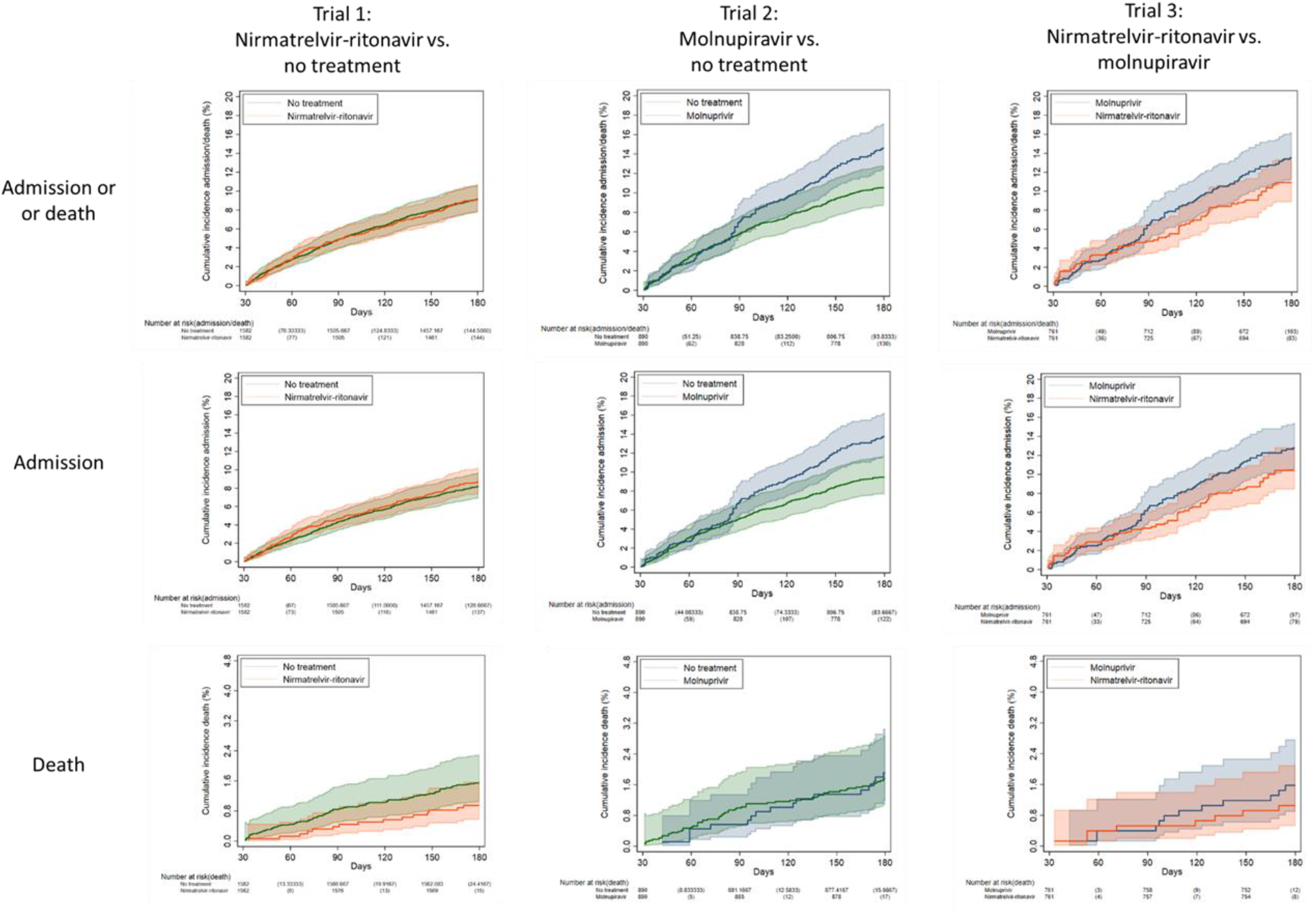
Kaplan-Meier curves comparing Veterans treated with nirmatrelvir-ritonavir versus their matched untreated counterparts (trial 1), molnupiravir versus their matched untreated counterparts (trial 2), and nirmatrelvir-ritonavir versus molnupiravir (trial 3) showing cumulative incidence (%) with 95% confidence intervals of any acute or long-term care admission or all-cause death, any acute or long-term care admission, and all-cause death 31-180 days after the index date among match groups alive at day 31.

**Table 3.**
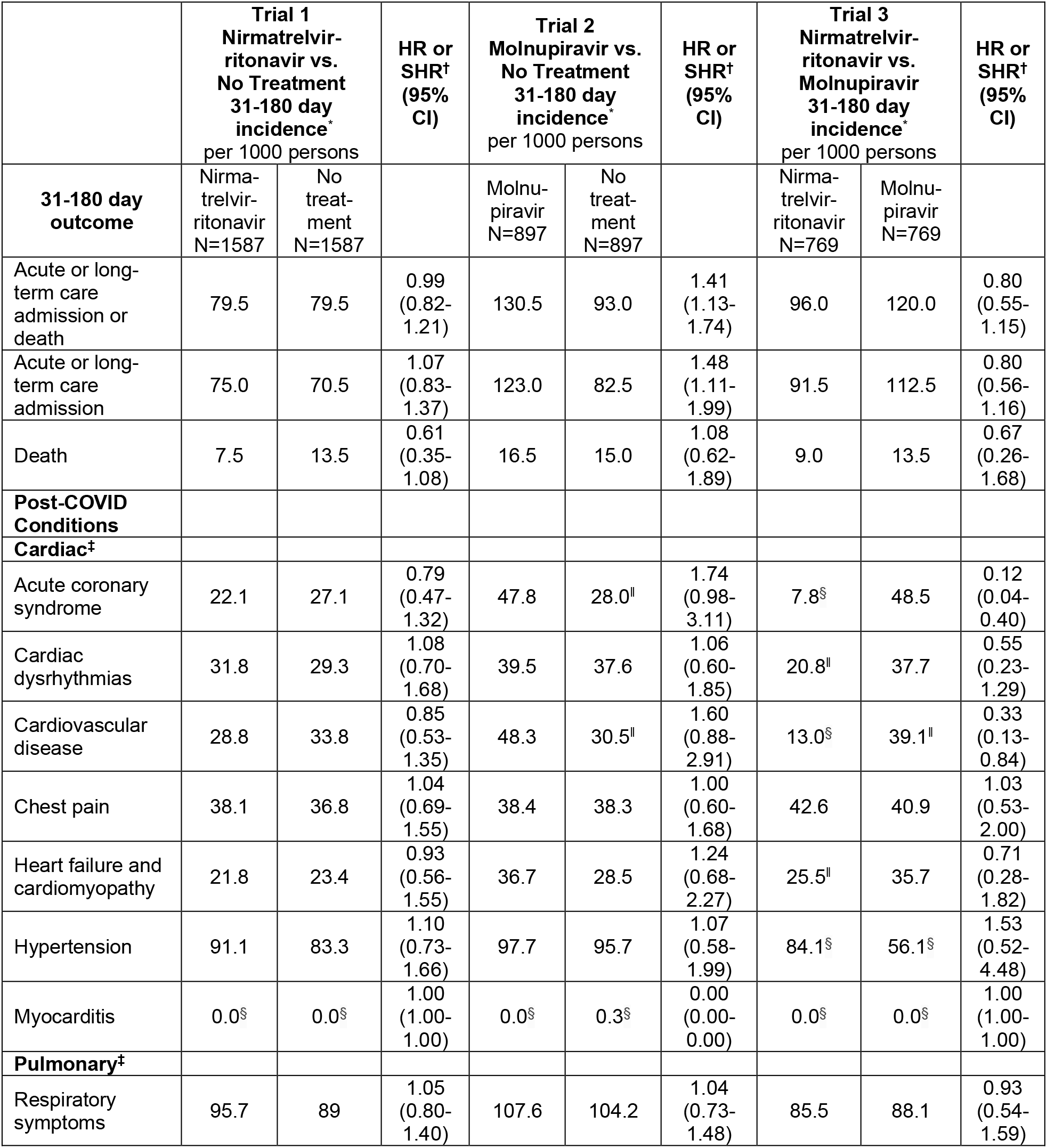
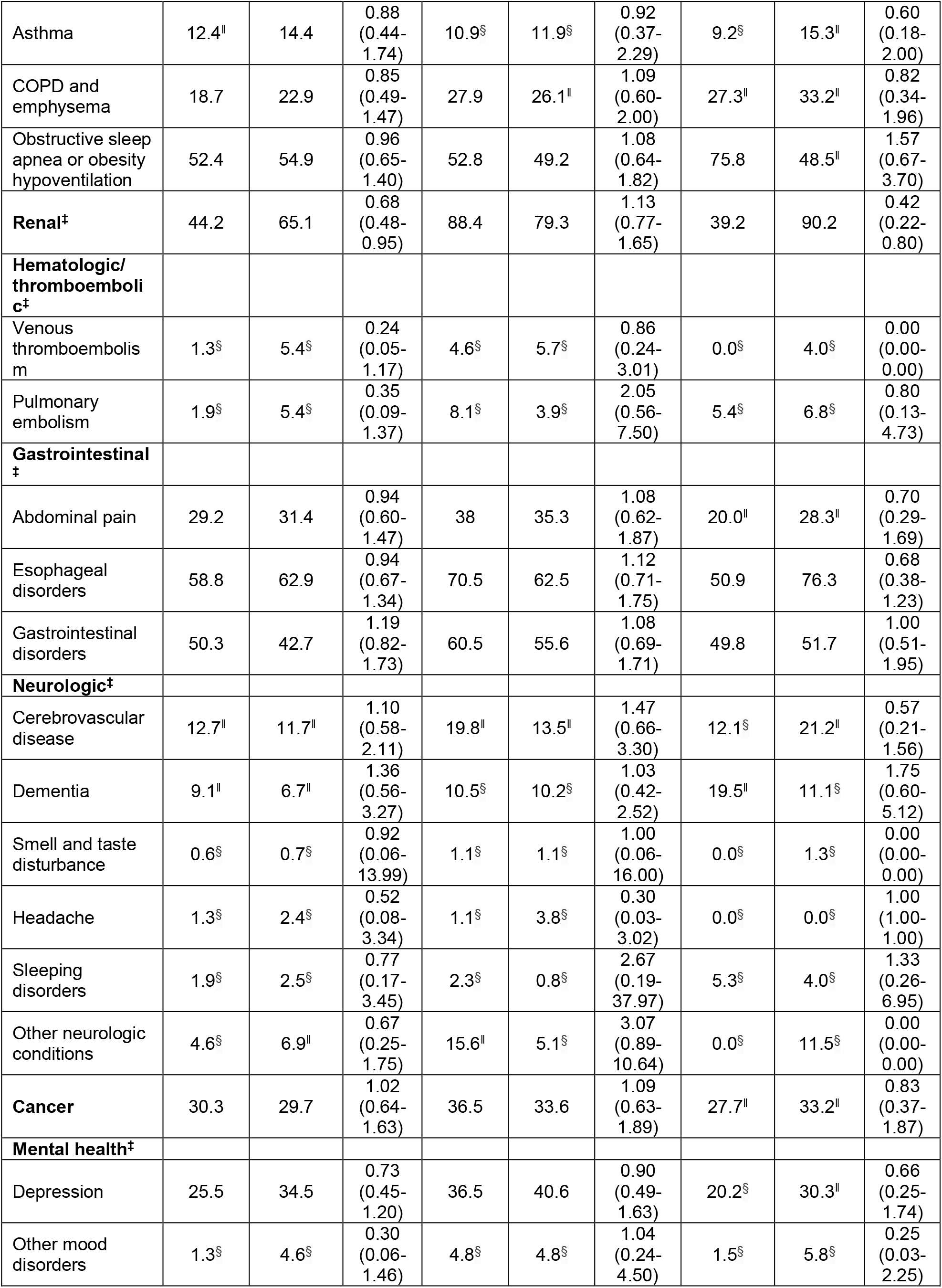
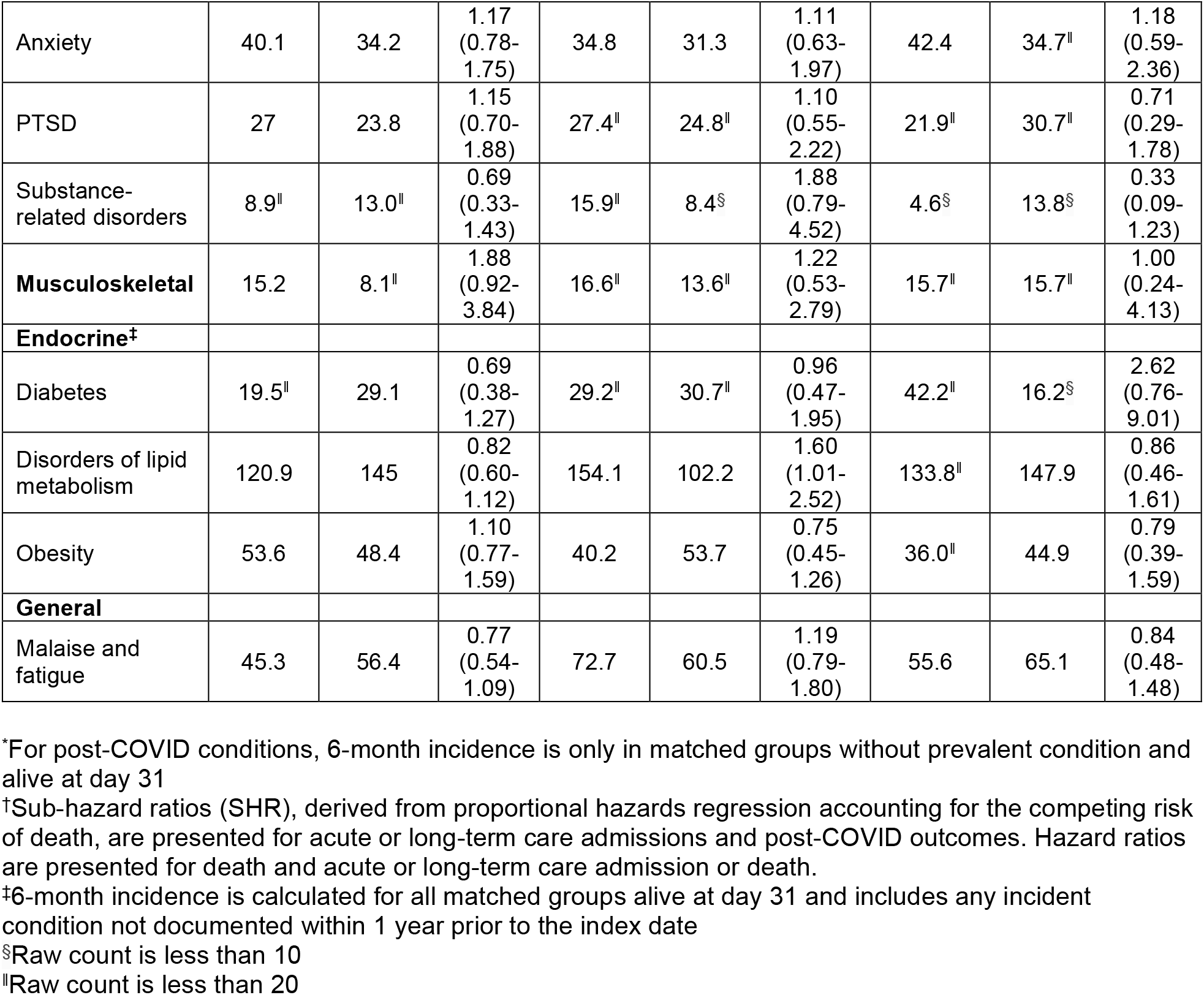
Comparison of matched groups in three emulated target trials of COVID-19 pharmacotherapy among Veterans who tested positive for SARS-CoV-2 from January 1, 2022 to February 28, 2022 with respect to cumulative 31-180 day incidence of hospitalization, death, and post-COVID conditions

|  | Trial 1<br>Nirmatrelvir-ritonavir vs.<br>No Treatment<br>31-180 day<br>incidence*<br>per 1000 persons |  | HR or<br>SHR†<br>(95%<br>CI) | Trial 2<br>Molnupiravir vs.<br>No Treatment<br>31-180 day<br>incidence*<br>per 1000 persons |  | HR or<br>SHR†<br>(95%<br>CI) | Trial 3<br>Nirmatrelvir-ritonavir vs.<br>Molnupiravir<br>31-180 day<br>incidence*<br>per 1000 persons |  | HR or<br>SHR†<br>(95%<br>CI) |
| --- | --- | --- | --- | --- | --- | --- | --- | --- | --- |
| 31-180 day<br>outcome | Nirma-<br>trelvir-<br>ritonavir<br>N=1587 | No<br>treat-<br>ment<br>N=1587 |  | Molnu-<br>piravir<br>N=897 | No<br>treat-<br>ment<br>N=897 |  | Nirma-<br>trelvir-<br>ritonavir<br>N=769 | Molnu-<br>piravir<br>N=769 |  |
| Acute or long-term care<br>admission or<br>death | 79.5 | 79.5 | 0.99<br>(0.82-1.21) | 130.5 | 93.0 | 1.41<br>(1.13-1.74) | 96.0 | 120.0 | 0.80<br>(0.55-1.15) |
| Acute or long-term care<br>admission | 75.0 | 70.5 | 1.07<br>(0.83-1.37) | 123.0 | 82.5 | 1.48<br>(1.11-1.99) | 91.5 | 112.5 | 0.80<br>(0.56-1.16) |
| Death | 7.5 | 13.5 | 0.61<br>(0.35-1.08) | 16.5 | 15.0 | 1.08<br>(0.62-1.89) | 9.0 | 13.5 | 0.67<br>(0.26-1.68) |
| **Post-COVID**<br>**Conditions** |  |  |  |  |  |  |  |  |  |
| **Cardiac‡** |  |  |  |  |  |  |  |  |  |
| Acute coronary<br>syndrome | 22.1 | 27.1 | 0.79<br>(0.47-1.32) | 47.8 | 28.0 | 1.74<br>(0.98-3.11) | 7.8<sup>§</sup> | 48.5 | 0.12<br>(0.04-0.40) |
| Cardiac<br>dysrhythmias | 31.8 | 29.3 | 1.08<br>(0.70-1.68) | 39.5 | 37.6 | 1.06<br>(0.60-1.85) | 20.8 | 37.7 | 0.55<br>(0.23-1.29) |
| Cardiovascular<br>disease | 28.8 | 33.8 | 0.85<br>(0.53-1.35) | 48.3 | 30.5 | 1.60<br>(0.88-2.91) | 13.0<sup>§</sup> | 39.1 | 0.33<br>(0.13-0.84) |
| Chest pain | 38.1 | 36.8 | 1.04<br>(0.69-1.55) | 38.4 | 38.3 | 1.00<br>(0.60-1.68) | 42.6 | 40.9 | 1.03<br>(0.53-2.00) |
| Heart failure and<br>cardiomyopathy | 21.8 | 23.4 | 0.93<br>(0.56-1.55) | 36.7 | 28.5 | 1.24<br>(0.68-2.27) | 25.5 | 35.7 | 0.71<br>(0.28-1.82) |
| Hypertension | 91.1 | 83.3 | 1.10<br>(0.73-1.66) | 97.7 | 95.7 | 1.07<br>(0.58-1.99) | 84.1<sup>§</sup> | 56.1<sup>§</sup> | 1.53<br>(0.52-4.48) |
| Myocarditis | 0.0<sup>§</sup> | 0.0<sup>§</sup> | 1.00<br>(1.00-1.00) | 0.0<sup>§</sup> | 0.3<sup>§</sup> | 0.00<br>(0.00-0.00) | 0.0<sup>§</sup> | 0.0<sup>§</sup> | 1.00<br>(1.00-1.00) |
| **Pulmonary‡** |  |  |  |  |  |  |  |  |  |
| Respiratory<br>symptoms | 95.7 | 89 | 1.05<br>(0.80-1.40) | 107.6 | 104.2 | 1.04<br>(0.73-1.48) | 85.5 | 88.1 | 0.93<br>(0.54-1.59) |
| Asthma | 12.4 | 14.4 | 0.88<br>(0.44-1.74) | 10.9<sup>§</sup> | 11.9<sup>§</sup> | 0.92<br>(0.37-2.29) | 9.2<sup>§</sup> | 15.3 | 0.60<br>(0.18-2.00) |
| COPD and emphysema | 18.7 | 22.9 | 0.85<br>(0.49-1.47) | 27.9 | 26.1 | 1.09<br>(0.60-2.00) | 27.3 | 33.2 | 0.82<br>(0.34-1.96) |
| Obstructive sleep apnea or obesity hypoventilation | 52.4 | 54.9 | 0.96<br>(0.65-1.40) | 52.8 | 49.2 | 1.08<br>(0.64-1.82) | 75.8 | 48.5 | 1.57<br>(0.67-3.70) |
| **Renal<sup>‡</sup>** | 44.2 | 65.1 | 0.68<br>(0.48-0.95) | 88.4 | 79.3 | 1.13<br>(0.77-1.65) | 39.2 | 90.2 | 0.42<br>(0.22-0.80) |
| **Hematologic/thromboembolic<sup>‡</sup>** |  |  |  |  |  |  |  |  |  |
| Venous thromboembolism | 1.3<sup>§</sup> | 5.4<sup>§</sup> | 0.24<br>(0.05-1.17) | 4.6<sup>§</sup> | 5.7<sup>§</sup> | 0.86<br>(0.24-3.01) | 0.0<sup>§</sup> | 4.0<sup>§</sup> | 0.00<br>(0.00-0.00) |
| Pulmonary embolism | 1.9<sup>§</sup> | 5.4<sup>§</sup> | 0.35<br>(0.09-1.37) | 8.1<sup>§</sup> | 3.9<sup>§</sup> | 2.05<br>(0.56-7.50) | 5.4<sup>§</sup> | 6.8<sup>§</sup> | 0.80<br>(0.13-4.73) |
| **Gastrointestinal<sup>‡</sup>** |  |  |  |  |  |  |  |  |  |
| Abdominal pain | 29.2 | 31.4 | 0.94<br>(0.60-1.47) | 38 | 35.3 | 1.08<br>(0.62-1.87) | 20.0 | 28.3 | 0.70<br>(0.29-1.69) |
| Esophageal disorders | 58.8 | 62.9 | 0.94<br>(0.67-1.34) | 70.5 | 62.5 | 1.12<br>(0.71-1.75) | 50.9 | 76.3 | 0.68<br>(0.38-1.23) |
| Gastrointestinal disorders | 50.3 | 42.7 | 1.19<br>(0.82-1.73) | 60.5 | 55.6 | 1.08<br>(0.69-1.71) | 49.8 | 51.7 | 1.00<br>(0.51-1.95) |
| **Neurologic<sup>‡</sup>** |  |  |  |  |  |  |  |  |  |
| Cerebrovascular disease | 12.7 | 11.7 | 1.10<br>(0.58-2.11) | 19.8 | 13.5 | 1.47<br>(0.66-3.30) | 12.1<sup>§</sup> | 21.2 | 0.57<br>(0.21-1.56) |
| Dementia | 9.1 | 6.7 | 1.36<br>(0.56-3.27) | 10.5<sup>§</sup> | 10.2<sup>§</sup> | 1.03<br>(0.42-2.52) | 19.5 | 11.1<sup>§</sup> | 1.75<br>(0.60-5.12) |
| Smell and taste disturbance | 0.6<sup>§</sup> | 0.7<sup>§</sup> | 0.92<br>(0.06-13.99) | 1.1<sup>§</sup> | 1.1<sup>§</sup> | 1.00<br>(0.06-16.00) | 0.0<sup>§</sup> | 1.3<sup>§</sup> | 0.00<br>(0.00-0.00) |
| Headache | 1.3<sup>§</sup> | 2.4<sup>§</sup> | 0.52<br>(0.08-3.34) | 1.1<sup>§</sup> | 3.8<sup>§</sup> | 0.30<br>(0.03-3.02) | 0.0<sup>§</sup> | 0.0<sup>§</sup> | 1.00<br>(1.00-1.00) |
| Sleeping disorders | 1.9<sup>§</sup> | 2.5<sup>§</sup> | 0.77<br>(0.17-3.45) | 2.3<sup>§</sup> | 0.8<sup>§</sup> | 2.67<br>(0.19-37.97) | 5.3<sup>§</sup> | 4.0<sup>§</sup> | 1.33<br>(0.26-6.95) |
| Other neurologic conditions | 4.6<sup>§</sup> | 6.9 | 0.67<br>(0.25-1.75) | 15.6 | 5.1<sup>§</sup> | 3.07<br>(0.89-10.64) | 0.0<sup>§</sup> | 11.5<sup>§</sup> | 0.00<br>(0.00-0.00) |
| **Cancer** | 30.3 | 29.7 | 1.02<br>(0.64-1.63) | 36.5 | 33.6 | 1.09<br>(0.63-1.89) | 27.7 | 33.2 | 0.83<br>(0.37-1.87) |
| **Mental health<sup>‡</sup>** |  |  |  |  |  |  |  |  |  |
| Depression | 25.5 | 34.5 | 0.73<br>(0.45-1.20) | 36.5 | 40.6 | 0.90<br>(0.49-1.63) | 20.2<sup>§</sup> | 30.3 | 0.66<br>(0.25-1.74) |
| Other mood disorders | 1.3<sup>§</sup> | 4.6<sup>§</sup> | 0.30<br>(0.06-1.46) | 4.8<sup>§</sup> | 4.8<sup>§</sup> | 1.04<br>(0.24-4.50) | 1.5<sup>§</sup> | 5.8<sup>§</sup> | 0.25<br>(0.03-2.25) |
| Anxiety | 40.1 | 34.2 | 1.17<br>(0.78-1.75) | 34.8 | 31.3 | 1.11<br>(0.63-1.97) | 42.4 | 34.7<sup>¶</sup> | 1.18<br>(0.59-2.36) |
| PTSD | 27 | 23.8 | 1.15<br>(0.70-1.88) | 27.4<sup>¶</sup> | 24.8<sup>¶</sup> | 1.10<br>(0.55-2.22) | 21.9<sup>¶</sup> | 30.7<sup>¶</sup> | 0.71<br>(0.29-1.78) |
| Substance-related disorders | 8.9<sup>¶</sup> | 13.0<sup>¶</sup> | 0.69<br>(0.33-1.43) | 15.9<sup>¶</sup> | 8.4<sup>§</sup> | 1.88<br>(0.79-4.52) | 4.6<sup>§</sup> | 13.8<sup>§</sup> | 0.33<br>(0.09-1.23) |
| **Musculoskeletal** | 15.2 | 8.1<sup>¶</sup> | 1.88<br>(0.92-3.84) | 16.6<sup>¶</sup> | 13.6<sup>¶</sup> | 1.22<br>(0.53-2.79) | 15.7<sup>¶</sup> | 15.7<sup>¶</sup> | 1.00<br>(0.24-4.13) |
| **Endocrine<sup>†</sup>** |  |  |  |  |  |  |  |  |  |
| Diabetes | 19.5<sup>¶</sup> | 29.1 | 0.69<br>(0.38-1.27) | 29.2<sup>¶</sup> | 30.7<sup>¶</sup> | 0.96<br>(0.47-1.95) | 42.2<sup>¶</sup> | 16.2<sup>§</sup> | 2.62<br>(0.76-9.01) |
| Disorders of lipid metabolism | 120.9 | 145 | 0.82<br>(0.60-1.12) | 154.1 | 102.2 | 1.60<br>(1.01-2.52) | 133.8<sup>¶</sup> | 147.9 | 0.86<br>(0.46-1.61) |
| Obesity | 53.6 | 48.4 | 1.10<br>(0.77-1.59) | 40.2 | 53.7 | 0.75<br>(0.45-1.26) | 36.0<sup>¶</sup> | 44.9 | 0.79<br>(0.39-1.59) |
| **General** |  |  |  |  |  |  |  |  |  |
| Malaise and fatigue | 45.3 | 56.4 | 0.77<br>(0.54-1.09) | 72.7 | 60.5 | 1.19<br>(0.79-1.80) | 55.6 | 65.1 | 0.84<br>(0.48-1.48) |

\*For post-COVID conditions, 6-month incidence is only in matched groups without prevalent condition and alive at day 31
<sup>†</sup>Sub-hazard ratios (SHR), derived from proportional hazards regression accounting for the competing risk of death, are presented for acute or long-term care admissions and post-COVID outcomes. Hazard ratios are presented for death and acute or long-term care admission or death.
<sup>‡</sup>6-month incidence is calculated for all matched groups alive at day 31 and includes any incident condition not documented within 1 year prior to the index date
<sup>§</sup>Raw count is less than 10
<sup>¶</sup>Raw count is less than 20

## DISCUSSION

In three target trial emulation studies performed among outpatient U.S. Veterans testing positive for SARS-CoV-2 during January and February 2022, nirmatrelvir-ritonavir was highly effective at preventing 30-day hospitalization or all-cause mortality, while risk reduction associated with molnupiravir was less substantial and limited to older groups. Receipt of nirmatrelvir-ritonavir was associated with a 47% lower risk in hospitalization or death through 30 days, including 79% lower risk of death and 34% lower risk of hospitalization. Overall, receipt of molnupiravir may have been associated with a lower risk of death through 30 days, and among Veterans aged 65 years and older, a lower combined risk of hospitalization or death. No additional statistically significant hospitalization or mortality benefit was observed from 31-180 days for either antiviral medication. Six-month incidence of renal conditions was lower among nirmatrelvir-ritonavir-treated participants, but benefit was not observed for any of the other post-COVID conditions nor among molnupiravir-treated participants.

While combined hospitalization and mortality benefit associated with nirmatrelvir-ritonavir was also observed in the Evaluation of Protease Inhibition for Covid-19 in High-Risk Patients (EPIC-HR) RCT (2), our estimated risk reduction was lower despite event rates of untreated groups in trial participants and our study being similar. EPIC-HR demonstrated an 89% relative reduction and 6 percentage point absolute reduction in 28-day COVID-19-related hospitalization or death while we observed a 47% lower risk of 30-day hospitalization or death corresponding with a 2.5 percentage point (25 per 1000 persons) reduction. Several factors may have accounted for differences in our findings, including differences in predominant circulating variants, COVID-19 vaccination, age of participants, and burden of underlying conditions. EPIC-HR was conducted during circulation of the Delta (B.1.617.2) variant in 2021, associated with more severe clinical outcomes (25), but by January 2022, Omicron (B.1.1.529) had become the main circulating variant. All participants in EPIC-HR were unvaccinated, whereas only 26% of participants in our study were unvaccinated; however, we did observe benefit among both vaccinated and unvaccinated Veterans in our study. Older age is associated with increased risk of severe COVID-related outcomes (26, 27), and our participants with median age of 65 years were nearly 20 years older than the median age in trial participants. We observed lower risk of hospitalization or death among Veterans aged 65 years and older but not younger groups aged 18-64 years. By comparison, although benefit was demonstrated for younger and older age groups in EPIC-HR, stronger benefit was seen among persons aged 65 years and older. Finally, Veterans in our study had notably higher prevalences of many underlying conditions associated with adverse outcomes (26), including diabetes, chronic kidney disease, and immunosuppression.

Several observational studies from Israel, Hong Kong, and the U.S. have demonstrated benefit associated with nirmatrelvir-ritonavir regarding short-term, severe COVID-19-related outcomes (3-8, 28). Very few studies, including EPIC-HR, were sufficiently powered to examine death (4, 7). The marked reduction in 30-day mortality seen in this study was similar to the effect size observed by Wong et al (7). Arbel et al. also reported a significant reduction in death due to COVID-19 but only among persons aged 65 years and older (4). To minimize common biases encountered in observational studies (9), we carefully defined time zero with regard to baseline eligibility and follow-up. Few COVID-19 pharmacotherapy studies minimized immortal time bias consistent with randomized trial design by matching the date of oral antiviral treatment among treated persons with the same number of days after the test-date for untreated persons (28). We were also careful to minimize selection bias by allowing as match-eligible untreated participants anyone who had not received COVID-19 pharmacotherapies through the date of oral antiviral treatment. Other studies that excluded untreated persons from the eligible pool based on treatment after the index date would have been subject to bias.

Although benefit from molnupiravir appeared less robust than nirmatrelvir-ritonavir, we found an overall absolute reduction in 30-day mortality as well as reduction in 30-day combined hospitalization and mortality among older Veterans. Evidence from clinical trials and observational studies of molnupiravir has also been mixed (7, 8, 29, 30). On one hand, the MOVe-OUT randomized trial demonstrated a small absolute risk reduction in 29-day hospitalization or death (29), and a reduction in mortality was also reported in one observational study (7). However, the PANORAMIC adaptive platform study did not find evidence of reduced 28-day hospitalization or death (30), and observational studies have not found evidence of reduced hospitalization (7, 8).

Our direct comparison of the two oral antivirals suggested a possible benefit of nirmatrelvir-ritonavir over molnupiravir. Although we had limited power to detect a small difference, there have been no head-to-head clinical trials nor observational studies in the U.S. to date (8). In the early months following FDA EUA of these two drugs in December 2021, a uniform preference for nirmatrelvir-ritonavir over molnupiravir as suggested by guidelines had not yet emerged in clinical practice (31-33), therefore providing a window of opportunity to conduct our observational comparative effectiveness study.

Finally, very little is known about the impact of oral antivirals on post-acute sequelae of SARS-CoV-2 infection (PASC). Our study is the first to provide information on 6-month outcomes, including molnupiravir-related outcomes, and incorporates VA Community care and CMS-Medicare data as well as comprehensive VA EHR data to strengthen the completeness of ascertained diagnoses. In contrast to other preliminary studies, we did not find clear evidence of reduced risk of PASC across other systems, including cardiovascular, pulmonary, endocrine, or neurologic conditions (34).

This study has several limitations. Eligibility for antiviral treatment of mild to moderate COVID-19 under FDA EUA requires symptom onset within 5 days, and we were not able to fully ascertain COVID-19-related symptom onset (31, 32). Although national surveillance is conducted by VA PBM to ensure eligibility among Veterans receiving treatment in the VHA, untreated comparators in this study may have included asymptomatic persons or symptomatic persons with delayed diagnosis beyond the eligible treatment window representing either more advanced disease or recovering illness. While the overall direction of potential bias may have favored oral antiviral agents compared with no treatment, we attempted to minimize this bias by requiring untreated matched persons to be alive and not hospitalized through the same number of days as the test-positive to treatment interval of the paired treated person. Furthermore, subgroup analysis still demonstrated benefit when stratified by presence or absence of any COVID-related symptoms within 30 days prior to testing positive. Second, this study was not designed to capture prior infections, which confer background immunity and may impact measured real-world effectiveness of antiviral treatments. However, the incidence of re-infection during January and February remained low (35). Third, capture of outpatient COVID-19 treatments and outcomes, particularly hospitalizations, may be incomplete. To address this, we restricted the study population to Veterans with a recent primary care visit who were more likely to seek care within the VHA system and integrated multiple data sources including CMS-Medicare to enhance ascertainment. Fourth, we could not verify whether Veterans prescribed antiviral medications completed treatment as recommended. Non-adherence may have biased estimates of effectiveness toward the null for comparisons with no treatment or favored molnupiravir over nirmatrelvir-ritonavir due to the distortion of taste associated with nirmatrelvir-ritonavir (2). Our findings reflect real-world conditions and do not strictly mirror intention-to-treat or per protocol analysis. Finally, only 3.8% of Veterans potentially eligible to receive outpatient anti-SARS-CoV-2 pharmacotherapies were estimated to receive any outpatient pharmacotherapy during this early period following FDA EUA of the oral antivirals (36).

Incidence of COVID-19 in the U.S. was also at its highest during January 2022 with 82% of ICU hospital beds occupied (37). Findings of this study may therefore not be generalizable to subsequent periods.

In conclusion, nirmatrelvir-ritonavir appears to be the most effective treatment for eligible persons with COVID-19 to reduce the risk of short-term, severe COVID-related outcomes.

Benefit may be greater in persons aged 65 years and older than younger adults. The role for molnupiravir may be more limited. Further studies are needed to clarify the long-term benefit of oral antivirals regarding incident post-COVID conditions.

## Data Availability

All data produced in the present work are contained in the manuscript.

## Disclaimer

The contents of this article do not represent the views of the US Department of Veterans Affairs or the US government.

## Additional Contributions

We thank the Biomedical Advanced Research and Development Authority (BARDA); US Food and Drug Administration (FDA); David Atkins, MD, Office of Research and Development (ORD), Department of Veterans Affairs (VA) Washington, DC; Matthew B. Goetz, MD, VA Greater Los Angeles Healthcare System and David Geffen School of Medicine at University of California, Los Angeles, CA; Rene LaFleur, MS, VA Cooperative Studies Program Clinical Epidemiology Research Center (CSP-CERC), VA Connecticut Healthcare System, West Haven, CT; William Lance, MPA, VA CSP-CERC, VA Connecticut Healthcare System, West Haven, CT; Alysia Maffucci, JD, VA CSP-CERC, VA Connecticut Healthcare System, West Haven, CT; Nicholas L. Smith, PhD, Seattle Epidemiologic Research and Information Center, Department of VA Office of Research and Development and Department of Epidemiology, University of Washington, Seattle, WA; and Xiao Qing Wang, MPH, Center for Clinical Management Research, VA Ann Arbor Healthcare System and Department of Internal Medicine, University of Michigan, Ann Arbor, MI for their support.

