## Supplemental Appendix for "Effectiveness of COVID-19 treatment with nirmatrelvir-ritonavir or molnupiravir among U.S. Veterans: target trial emulation studies with one-month and six-month outcomes"

### Contents

Contents ... 2

A. SUPPLEMENTAL METHODS ... 4

Supplemental Table 1. Specification and emulation of three target randomized controlled trials of COVID-19 antiviral agents among non-hospitalized adult VA enrollees who tested positive for SARS-CoV-2 from January 1-February 28, 2022, using observational data from the VHA EHR ... 4

Departures from the protocol for the comparison of nirmatrelvir-ritonavir versus molnupiravir 8

Eligibility criteria and study population ... 8

Supplemental Table 2. Risk factors for progression to severe COVID-19 according to the CDC(4) ... 8

Supplemental Table 3. Immunosuppressive and cancer medications* ... 9

Exclusions for nirmatrelvir-ritonavir comparisons ... 10

Supplemental Table 4. List of medications contraindicated with use of nirmatrelvir-ritonavir* ... 10

Cohort matching ... 11

Supplemental Table 5. National Institutes of Health (NIH) tiers of prioritization for anti-SARS-CoV-2 therapies* ... 11

COVID-19 Vaccination Status ... 11

Supplemental Table 6. Characteristics included in propensity score matching models for COVID-19 treatment* ... 12

Supplemental Table 7. List of underlying conditions totaled for inclusion in propensity score matching models for COVID-19 treatment* ... 13

Additional exclusions applied during matching ... 14

Sample size and power calculations ... 14

Supplemental Table 8. List of conditions evaluated as potential post-COVID incident conditions from day 31 to 180 after the index date* ... 14

B. SUPPLEMENTAL RESULTS ... 16

Supplemental Table 9. Additional persons excluded from the match-eligible cohort during matching ... 16

Supplemental Table 10. Baseline demographic and clinical characteristics of Veterans who tested positive for SARS-CoV-2 in the VA healthcare system from January 1, 2022 to February 28, 2022, who fulfilled eligibility criteria in three emulated target trials of COVID-19 antiviral effectiveness ... 17

Supplemental Figure 1. Distribution of baseline characteristics and propensity scores in persons who were treated with nirmatrelvir-ritonavir and their comparators receiving no COVID-19 antiviral or monoclonal antibody treatment, before and after matching. ... 20

### A. SUPPLEMENTAL METHODS

**Supplemental Table 1. Specification and emulation of three target randomized controlled trials of COVID-19 antiviral agents among non-hospitalized adult VA enrollees who tested positive for SARS-CoV-2 from January 1-February 28, 2022, using observational data from the VHA EHR**

Trial 1: Nirmatrelvir-ritonavir versus no outpatient SARS-CoV-2 treatment*
Trial 2. Molnupiravir versus no outpatient SARS-CoV-2 treatment*
Trial 3. Nirmatrelvir-ritonavir versus molnupiravir

| Target Trials Specification | Target Trials Emulation |
|---|---|
| **Eligibility Criteria for All Three Trials** | |
| Aged ≥18 years at the time of positive SARS-CoV-2 test performed January 1, 2022-February 28, 2022 | Same |
| VA enrollees (excludes VA employees who are not enrollees) | Same |
| Followed by a primary care provider in the VA healthcare system, defined as having a primary care outpatient encounter in the preceding 18 months | Same |
| Documented first positive laboratory-based SARS-CoV-2 NAAT or antigen test in a respiratory specimen performed between January 1-February 28, 2022. Patients with reinfection during this period (i.e., those who also have a documented positive SARS-CoV-2 NAAT or antigen test before January 1, 2022) were not included. | Same<br><br>We identified all VA enrollees who had a first positive laboratory-based SARS-CoV-2 NAAT or antigen test using the VA COVID-19 National Surveillance Tool (NST) as documented in CSDR. This includes all patients tested within the VA as well as patients with such tests performed outside the VA but documented in VA records. Tests performed outside the VA are identified by methods including NLP and confirmed by manual review of the EHR before being documented in the NST. |
| Symptomatic infection (≥1 symptoms) at the time of diagnosis (1, 2) | Not included.<br><br>CSDR captures 15 pre-specified COVID-19-related symptoms documented in the EHR in the 30 days prior to the first positive SARS-CoV-2 test. Because ascertainment of symptoms is incomplete and since CSDR does not distinguish whether symptoms were present at the time of testing positive, we did not require presence of ≥1 symptoms. Presence or absence of symptoms were included in propensity score matching instead. |

| | |
|---|---|
| Not hospitalized on or before the date of the positive SARS-CoV-2 test (test-positive date) and not hospitalized on or before the date of randomized assignment to nirmatrelvir-ritonavir, molnupiravir, or no treatment. | Same<br><br>Participants were not hospitalized on or within 7 days before test-positive date.<br><br>Participants receiving nirmatrelvir-ritonavir or molnupiravir were not hospitalized on or before the date of treatment initiation.<br><br>Matched untreated participants were not hospitalized on or before their assigned index date (which was the same number of days from the test positive date as the matched treated patient, see below) |
| Did not initiate therapy before the test-positive date | Same |
| Persons treated with nirmatrelvir-ritonavir or molnupiravir did not receive more than one outpatient COVID-19 therapy within 7 days. Untreated persons did not receive any other outpatient COVID-19 therapy on or prior to the matched antiviral treatment date. | Same<br><br>Persons receiving nirmatrelvir-ritonavir or molnupiravir were not treated with other outpatient COVID-19 treatments (nirmatrelvir-ritonavir, molnupiravir, sotrovimab, remdesivir) on or prior to the antiviral treatment date. Untreated persons did not receive any other outpatient COVID-19 therapy on or prior to the matched antiviral treatment date. |
| Having ≥1 risk factors for progression to severe COVID-19 by the FDA EUA/CDC criteria (**Supplemental Table 3**) | Same |
| **Additional Eligibility Criteria for Participants in Trials Including a Nirmatrelvir-ritonavir Arm** | |
| Persons with any of the following were excluded:<br>• Moderate or severe liver disease<br>• Advanced renal impairment (CKD IV or V, on dialysis, or eGFR<30)<br>• Prescription contraindicated medications per FDA in the 90 days prior to test-positive date (**Supplemental Table 4**) | Same |
| **Additional Eligibility Criteria for Participants in Trials Including a Molnupiravir Arm** | |
| Pregnant persons were excluded | Same |
| **Treatment Strategies** | |
| Randomized to treatment with one of the following within 5 days of symptom onset:<br>• Nirmatrelvir-ritonavir<br>• Molnupiravir<br>• No treatment | We determined the date of treatment with nirmatrelvir-ritonavir or molnupiravir from pharmacy fill date of the medication.<br><br>Since symptom onset date could not be ascertained in the EHR, we used treatment |

<table>
<tr><td></td><td>within 10 days of the test-positive date rather within 5 days of symptom onset. In clinical practice, adherence to EUA criteria (treatment within 5 days of symptom onset) is closely monitored and enforced by PBM.</td></tr>
<tr><td colspan="2">Treatment Assignment</td></tr>
<tr><td>In each of the three RCTs eligible participants are randomly assigned to each of the two treatment options:<br>• Nirmatrelvir-ritonavir vs no treatment<br>• Molnupiravir vs. no treatment<br>• Nirmatrelvir-ritonavir vs. molnupiravir</td><td>Sequential matching:<br>Exact-matching<br>We first exact-matched each eligible participant who received nirmatrelvir-ritonavir or molnupiravir to all eligible participants who did not receive any treatment using four factors:<br>• NIH tier of prioritization for anti-SARS-CoV-2 therapies (Supplementary Table 4)<br>• VISN, the 19 geographical administrative regions of the VA<br>• Calendar time, centered +/- 7 days around the test-positive date of the matched comparator<br>For the comparison of nirmatrelvir-ritonavir versus molnupiravir, additional exact-matching based on the interval from test-positive date to treatment date (0-1 days vs. ≥2 days) was used.<br><br>Propensity-score matching<br>After exact-matching by these four factors, we performed an additional propensity score matching step ultimately aiming to identify the best-matching comparator. We used matching with replacement in a 1:k variable ratio, where k varied based on the number of propensity score ties. We included all ties to avoid imbalance due to random pruning.<br>The characteristics included in the propensity score logistic regression model are shown in Supplemental Table 6.</td></tr>
<tr><td colspan="2">Outcomes</td></tr>
<tr><td><u>Primary Outcome</u>:<br>Hospitalization lasting >24 hours for any cause or death through day 30<br><br><u>Secondary Outcomes</u>:<br>Intensive care Unit (ICU) admission through day 30<br>Mechanical ventilation through day 30</td><td>Same<br><br>Hospitalization, ICU admission and mechanical ventilation were determined using a combination of VA/CDW, Community Care and CMS-Medicare data.<br><br>Post-COVID Conditions were determined using VA/CDW data</td></tr>
</table>

| | |
|---|---|
| Hospital and outpatient encounter-free days (urgent care/office/phone/telehealth visit) through 90 days post-treatment<br><br><u>Post-COVID Conditions:</u><br>Incidence at 31-180 days of the following conditions:<br>• Cardiac, pulmonary, renal, GI, neurologic, cancer, mental health, musculoskeletal, endocrine<br>• All-cause mortality | |
| **Follow-up** | |
| For each person, follow-up started on the day of randomization for each of the three target trials and continued until day 30 after treatment for short-term outcomes or from day 31-180 for post-COVID-19 condition outcomes. | Same<br><br>For comparisons with untreated patients, an index date was assigned to untreated patients which was the same interval from the test-positive date. Untreated persons had to fulfill enrollment criteria as of that index date (e.g., a person treated on day 3 after testing positive was matched to a person who was untreated, alive, and not hospitalized 3 days after testing positive). This study design ensured that dates of eligibility determination, treatment, and follow-up initiation were the same (**Supplemental Figure 1**). |
| **Causal Contrasts** | |
| Intention-to-treat (ITT) effect | Observational analogue of the per-protocol effect |
| Per-protocol effect (i.e., the effect if all individuals had received their assigned treatment) | Observational analogue of the per-protocol effect |
| **Statistical Analysis** | |
| 30-day risk, risk difference and risk ratios for the primary outcome.<br>31-180-day incidence of any acute or long-term care admission or all-cause death<br>31-180-day incidence of post-COVID conditions | Same |

[*]Includes treatments available in the VA during the study period (nirmatrelvir-ritonavir, molnupiravir, sotrovimab, and outpatient remdesivir).

Abbreviations: ATSDR, Agency for Toxic Substances and Disease Registry; CAN, Care Assessment Needs; CDC, Centers for Disease Control and Prevention; CLC, Community Living Center; CDW, Corporate Data Warehouse; CSDR, COVID-19 Shared Data Resource; eGFR, estimated glomerular filtration rate; EHR, electronic health records; EUA, emergency use authorization; FDA, Food and Drug Administration; ICU, Intensive Care Unit; ITT, Intention-to-treat; NAAT, nucleic acid amplification test; NIH, National Institutes of Health; NLP, Natural

Language Processing; NST, National Surveillance Tool; PASC, Post-acute Sequelae of COVID-19; PBM, Pharmacy Benefits Management Services; PCP, Primary Care Provider; SVI, Social Vulnerability Index; VAMC, VA Medical Center; VISN, VA Integrated Services Network

**Departures from the protocol for the comparison of nirmatrelvir-ritonavir versus molnupiravir**

Due to insufficient matching for the comparison of nirmatrelvir-ritonavir with molnupiravir, we expanded the calendar time match window from +/- 7 days to +/- 10 days around the test-positive date of the matched comparator in the exact-matching step. For propensity-score matching, we used VA region rather than VISN(3) and did not include chronic kidney disease since differences in nirmatrelvir-ritonavir prescribing practices for persons with eGFR 30-59 ml/min (by either dose-adjusting nirmatrelvir-ritonavir or considering an alternative agent) were anticipated to result in difficulty matching on this variable.

**Eligibility criteria and study population**

Participants in all three emulated trials were limited to test-positive VA enrollees. We excluded persons who received nirmatrelvir-ritonavir or molnupiravir before or after 10 days following the test-positive date, determined to be a reasonable proxy treatment window relative to the 5-day window from symptom onset per FDA EUA. While it is difficult to accurately ascertain symptom onset from the EHR, VA Pharmacy Benefits Management Services (PBM) monitors and enforces adherence to EUA. We also limited eligibility to Veterans with at least one risk factor for progression to severe COVID-19, including hospitalization or death, according to the Centers for Disease Control and Prevention (CDC)(4) as shown in **Supplemental Table 2** below.

**Supplemental Table 2. Risk factors for progression to severe COVID-19 according to the CDC(4)**

| |
|---|
| **Age ≥ 65 years*** |
| **Underlying Medical Conditions** |
| Cancer |
| Cardiovascular disease including cardiomyopathy, chronic rheumatic heart disease, congestive heart failure, coronary artery disease, hypertension, myocardial infarction, peripheral artery disease, pulmonary heart disease |
| Chronic kidney disease including dialysis |
| Chronic liver disease including chronic hepatitis and cirrhosis |
| Chronic lung disease including asthma, chronic obstructive pulmonary disease, emphysema, pulmonary fibrosis |
| Chronic neurologic conditions including epilepsy, multiple sclerosis, and Parkinson's disease |
| Dementia |
| PTSD |
| Diabetes |
| HIV |
| Immunosuppressive medications or cancer therapies (see Supplementary Table 3) |
| Mental health conditions including bipolar disorder, major depressive disorder, post-traumatic stress disorder, and schizophrenia |
| Overweight (body mass index 25 to <30 kg/m$^2$) or obese (body mass index ≥30 kg/m$^2$) or |
| Pregnancy |
| Sickle cell disease |

| |
|---|
| Stroke or cerebrovascular disease |
| Thalassemia |
| **Current or former tobacco use** |
| **Substance use** |
| Alcohol dependence |
| Non-alcohol substance dependence |

*Age at the time of positive SARS-CoV-2 test or underlying conditions, tobacco, or substance use documented in the 2 years prior to positive SARS-CoV-2 test.

**Supplemental Table 3. Immunosuppressive and cancer medications***

| |
|---|
| ABATACEPT, ABEMACICLIB, ABRAXANE, ACALABRUTINIB, ADALIMUMAB, AFATINIB, ALDESLEUKIN, ALECTINIB, ALEMTUZUMAB,† ALPELISIB, ANAKINRA, ANTI-THYMOCYTE GLOBULIN, † APREMILAST, APSPARAGINASE, ARSENIC TRIOXIDE, ASCIMINIB, ASPARAGINASE, ATEZOLIZUMAB, AVACOPAN, AVAPRITINIB, AVELUMAB, AXITINIB, AZACITIDINE, AZATHIOPRINE, BARICITINIB, BASILIXIMAB, BELANTAMAB, BELATACEPT, BELIMUMAB, BELINOSTAT, BELUMOSUDIL, BELZUTIFAN, BENDAMUSTINE, BENRALIZUMAB, BEVACIZUMAB, BEXAROTENE, BINIMETINIB, BINMETINIB, BLEOMYCIN, BLINATUMOMAB, BORTEZOMIB, BOSUTINIB, BRENTUXIMAB, BRIGATINIB, BRODALUMAB, BUDESONIDE, BUSULFAN, CABAZITAXEL, CABOZANTINIB, CANAKINUMAB, CAPECITABINE, CAPMATINIB, CARBOPLATIN, CARFILZOMIB, CARMUSTINE, CEMIPLIMAB, CERITINIB, CERTINIB, CERTOLIZUMAB, CETUXIMAB, CHLORAMBUCIL, CISPLATIN, CISPLATINUM, CLADRIBINE, CLOFARABINE, COBIMETINIB, COPANLISIB, COPAXONE, CRIZOTINIB, CYCLOPHOSPHAMIDE, CYCLOSPORINE, CYTARABINE, DABRAFENIB, DACARBAZINE, DACOMITINIB, DACTINOMYCIN, DARATUMUMAB, DASATINIB, DAUNORUBICIN, DENOSUMAB, DEXAMETHASONE, DIMETHYL FUMARATE, DOCETAXEL, DOXORUBICIN, DUPILUMAB, DURVALUMAB, DUVELISIB, ECULIZUMAB, ELOTUZUMAB, ENASIDENIB, ENCORAFENIB, ENFORTUMAB, ENTRECTINIB, ENZALUTAMIDE, EPIRUBICIN, ERDAFITINIB, ERIBULIN, ERLOTINIB, ESTRAMUSTINE, ETANERCEPT, ETOPOSIDE, EVEROLIMUS, FATUMUMAB, FINGOLIMOD, FLUDARABINE, FLUOROURACIL, FLUTAMIDE, GEFITINIB, GEMCITABINE, GEMTUZUMAB, GENGRAF, GILTERITINIB, GLASDEGIB, GLATIRAMER, GLATIRAMIR ACETATE, GOLIMUMAB, GUSELKUMAB, HERCEPTIN, HYDROCORTISONE, HYDROXYCHLOROQUINE, HYDROXYUREA, IBRUTINIB, IDARUBICIN, IDELALISIB, IFOSFAMIDE, IMATINIB, INFIGRATINIB, INFLECTRA, INFLIXIMAB, INOTUZUMAB, INTERFERON, INVIBRUTINIB, IPILIMUMAB, IRINOTECAN, ISATUXIMAB, IVOSIDENIB, IXAZOMIB, IXEKIZUMAB, LAPATINIB, LAROTRECTINIB, LEFLUNOMIDE, LENALIDOMIDE, LENVATINIB, LETROZOLE, LOMUSTINE, LONCASTUXIMAB, LURBINECTEDIN, MARGETUXIMAB, MECHLORETHAMINE, MELPHALAN, MEPOLIZUMAB, MERCAPTOPURINE, METHOTREXATE, METHYLPREDNISOLONE, MIDOSTAURIN, MITOMYCIN, MITOXANTRONE, MOGAMULIZUMAB, MYCOPHENOLATE, MYCOPHENOLIC ACID, NATALIZUMAB, NELARABINE, NERATINIB, NILOTINIB, NILUTAMIDE, NIRAPARIB, NIVOLUMAB, OBINUTUZUMAB, OCRELIZUMAB,† OFATUMUMAB,† OLAPARIB, OSIMERTINIB, OXALIPLATIN, PACLITAXEL, PALBOCICLIB, PANITUMUMAB, PANOBINOSTAT, PAZOPANIB, PEGASPARGASE, PEGINTERFERON, PEMBROLIZUMAB, PEMETREXED, PEMIGATINIB, PENTOSTATIN, PERTUZUMAB, PEXIDARTINIB, PIMECROLIMUS, POLATUZUMAB, POMALIDOMIDE, PONATINIB, PRALATREXATE, PREDNISOLONE, PREDNISONE, PROCARBAZINE, RAMUCIRUMAB, RASBURICASE, RAVULIZUMAB, REGORAFENIB, RENFLEXIS, RESLIZUMAB, RIBOCICLIB, RILONACEPT, |

| |
|---|
| RIPRETINIB, RISANKIZUMAB, RISKANIKIZUMAB, RITUXIMAB,[†] ROMIDEPSIN, ROPEGINTERFERON, RUCAPARIB, RUXOLITINIB, SACITUZUMAB, SARILUMAB, SATRALIZUMAB, SECUKINUMAB, SELINEXOR, SELPERCATINIB, SILTUXIMAB, SIPONIMOD, SIPULEUCEL, SIROLIMUS, SORAFENIB, SULFASALAZINE, SUNITINIB, SUTIMLIMAB, TACROLIMUS, TAFASITAMAB, TAGRAXOFUSP, TALAZOPARIB, TAZEMETOSTAT, TEMOZOLOMIDE, TEMSIROLIMUS, TERIFLUNOMIDE, THALIDOMIDE, THIOGUANINE, THIOPHOSPHORAMIDE, THIOTEPA, TILDRAKIZUMAB, TIPIRACIL, TIVOZANIB, TOCILIZUMAB, TOFACITINIB, TRABECTEDIN, TRAMETINIB, TRASTUZUMAB, TRIFLURIDINE, TUCATINIB, UMBRALISIB, UPADACITINIB, USTEKINUMAB, VALRUBICIN, VANDETANIB, VEDOLIZUMAB, VENETOCLAX, VINBLASTINE, VINCRISTINE, VINORELBINE, VORINOSTAT, ZANUBRUTINIB |

[*]Prescriptions filled within 90 days of test-positive date unless otherwise indicated
[†]Filled within one year of test-positive date

**Exclusions for nirmatrelvir-ritonavir comparisons**
For comparisons involving nirmatrelvir-ritonavir, we excluded persons with advanced renal impairment, defined as any of following:

- Estimated glomerular filtration rate (eGFR) less than 30 milliliters per minute based on labs obtained in the six months prior to positive SARS-CoV-2 test (test date) and calculated using Chronic Kidney Disease Epidemiology Collaboration (CKD-EPI) 2021.
- Diagnosis of stage VI or V chronic kidney disease or end stage renal disease based on specific ICD10 codes (N18.4, N18.5, N18.6) recorded as least once in the two years prior to test date.
- Dialysis at test date.

We also excluded persons with moderate or severe liver disease documented in the two years prior to the test date as captured by VA's COVID-19 Shared Data Resource (CSDR). For comparisons involving nirmatrelvir-ritonavir, we excluded persons with a prescription for contraindicated medications filled within 90 days prior to the test date as shown in the table below.(5)

**Supplemental Table 4. List of medications contraindicated with use of nirmatrelvir-ritonavir[*]**

| | | |
|---|---|---|
| Alfuzosin | Ivabradine | Ranolazine |
| Amiodarone | Ivacaftor/lumacaftor | Rifampicin |
| Apalutamide | Lomitapide | Rifampin |
| Carbamazepine | Lurasidone | Sildenafil |
| Colchicine | Methylergonovine | Silodosin |
| Dihydroergotamine | Midazolam | St. John's Wort |
| Dronedarone | Naloxegol | Tolvaptan |
| Eletriptan | Phenobarbital | Triazolam |
| Eplerenone | Phenytoin | Ubrogepant |
| Ergotamine | Pimozide | Voclosporin |
| Flibanserin | Primadone | |
| Finerenone | Propafenone | |
| Flecainide | Quinidine | |

*List modified from the Food and Drug Administration patient eligibility screening checklist tool for prescribers. Lovastatin and simvastatin have been excluded from this list as they can be held during and for up to 5 days after treatment with nirmatrelvir-ritonavir.

**Cohort matching**

We used two matching steps, exact and propensity score matching to achieve balance of covariates between comparator groups and reduce confounding. NIH tiers of prioritization used for exact-matching are shown in the Table below:

**Supplemental Table 5. National Institutes of Health (NIH) tiers of prioritization for anti-SARS-CoV-2 therapies***

| NIH Tier | Risk Group Definition† |
|---|---|
| 1 | - Receipt of immunosuppressive or cancer medications‡ *or*<br>- HIV with most recent absolute CD4 count ≤2 years is ≤200 cells/mm³ *or*<br>- Unvaccinated§ and age ≥75 years *or*<br>- Unvaccinated, age 65-74 years, and ≥1 risk factor for severe COVID-19‖ |
| 2 | - Unvaccinated, age 65-74 years, and no risk factors for severe COVID-19 *or*<br>- Unvaccinated, age <65 years, and ≥1 risk factor for severe COVID-19 |
| 3 | - Vaccinated§ and age ≥75 years *or*<br>- Vaccinated, age 65-74 years and ≥1 risk factor for severe COVID-19 |
| 4 | - Vaccinated, age 65-74 years, and no risk factors for severe COVID-19 *or*<br>- Vaccinated, age <65 years, and ≥1 risk factor for severe COVID-19 |
| Other | - Vaccination status not determined (individuals not included in Tier 1) |

*Per NIH definitions: https://www.covid19treatmentguidelines.nih.gov/overview/prioritization-of-therapeutics/?msclkid=9db67596cf2311ec8b6f159cc2c59087
†All factors determined with reference to positive SARS-CoV-2 test
‡As defined in Supplemental Table 3
§As defined in under COVID-19-Vaccination Status. To categorize vaccination as a binary variable for the National Institutes of Health (NIH) tiers of prioritization for anti-SARS-CoV-2 therapies, we considered unvaccinated and partially vaccinated Veterans as 'unvaccinated' for tiers 1 and 2 and fully vaccinated and boosted as 'vaccinated'
‖As defined in Supplemental Table 2

**COVID-19 Vaccination Status**

We aggregated all administered vaccine doses documented in VA-CDW, CMS-Medicare and VA Community Care data. Vaccine records with service dates prior to December 11, 2020, the earliest date of EUA for COVID-19 vaccination in the United States, were excluded. To ensure that vaccine doses documented in more than one source were not counted more than once, after combining records from all sources, we treated 2 vaccine doses as duplicates if they were documented within 7 days of each other.

We included Moderna, Pfizer-BioNTech, and Janssen vaccine types, which were approved in the United States during the period of study and comprised most of all vaccine types. To allow for complete categorization of vaccination, we also included Novavax (authorized after the end of this study period). Vaccine doses of unknown or other type (e.g., Oxford-AstraZeneca) were categorized as other.

Non-immunocompromised (6)

1. Veterans were considered unvaccinated if they did not receive any COVID-19 vaccine or received a vaccine dose other than Janssen less than 14 days prior to the first positive SARS-CoV-2 test (test date).
2. Partial vaccination was indicated by receipt of a single mRNA dose (Pfizer-BioNTech or Moderna) or a single Novavax dose alone or in combination with another vaccine other than Janssen <14 days prior to the index date or a Janssen (Johnson & Johnson) dose <14 days before the test date.
3. Primary vaccination was indicated by receipt of 2 doses of any mRNA or Novavax vaccine or a single dose of Janssen ≥14 days before the test date.
4. Booster vaccination was indicated by any primary regimen above, followed by an additional dose(s) of mRNA, Janssen, or Novavax vaccine ≥7 days before the test date.
5. Other was indicated by any vaccination not captured above.

Immunocompromised (7)

Veterans were considered immunocompromised if they had recently received immunosuppressive or cancer medications as described in Supplementary Table 3).

1. Veterans were considered unvaccinated if they did not receive any COVID-19 vaccine or received a vaccine dose other than Janssen less than 14 days prior to the test date.
2. Partial vaccination was indicated by receipt of 2 doses of an mRNA vaccine, a single dose of Janssen, or a single dose of Novavax. It was also indicated by receipt of 3 doses of an mRNA vaccine, a single dose of Janssen followed by a single dose of an mRNA vaccine, or 2 doses of Novavax <7 days before the test date.
3. Primary vaccination was indicated by receipt of 3 doses of an mRNA vaccine, a single dose of Janssen followed by a single dose of an mRNA vaccine, or 2 doses of Novavax ≥7 days before the test date.
4. Booster vaccination was indicated by any primary regimen above, followed by an additional dose(s) of mRNA, Janssen, or Novavax vaccine ≥7 days before the test date.
5. Other was indicated by any vaccination not captured above.

The Table below describes factors included in a propensity logistic regression model.

**Supplemental Table 6. Characteristics included in propensity score matching models for COVID-19 treatment***

| |
|---|
| **Calendar week of test** |
| **Demographic** |
| Age |
| Sex |
| Race/ethnicity |
| **Geographic** |
| Urban/rural residence based in RUCA codes (rurality)(8) |
| Area Deprivation Index (ADI)(9) |
| **Substance use** |
| Tobacco |
| Alcohol |

| |
|---|
| Other substances |
| **Presence of any COVID-19 symptoms documented in the 30 days prior to the test-positive date†** |
| **COVID-19 vaccination** |
| None, partial, primary, booster and other |
| Time since completion of last vaccine dose |
| **Underlying conditions** |
| BMI (<18.5, 18.5-24.9, 25-29.9, 30-34.9, 35-39.9, ≥40 kg/m$^2$) |
| Number of high-risk conditions for progression to severe COVID-19 (Supplemental Table 7) |
| Chronic kidney disease‡ |
| Diabetes |
| Immunosuppressive and cancer medications (Supplemental Table 3) |
| Cardiovascular disease |
| Chronic lung disease |
| Dementia |
| Care Assessment Needs (CAN) score |
| **Healthcare utilization: number of clinical encounters** |

*Unless otherwise specified, factors were ascertained as of the index date. Urban/rural residence and ADI were determined using ZIP codes for Veteran's most recent place of residence in the one year prior to index date. Substance use and underlying conditions were ascertained based on documentation in the two years prior to index date. Number of outpatient visits of any type were counted in the one year prior to index date.
†Any of 15 pre-specified COVID-19-related symptoms present on the day of positive SARS-CoV-2 test or within the preceding 30 days.
‡Not included in molnupiravir vs nirmaltrevir trial.

**Supplemental Table 7. List of underlying conditions totaled for inclusion in propensity score matching models for COVID-19 treatment***

| |
|---|
| Any immunosuppressive or cancer medications (Supplemental Table 3) |
| Acute myocardial infarction or coronary atherosclerosis |
| Asthma |
| Cancer |
| Cardiomyopathy or heart failure |
| Cerebrovascular disease including stroke |
| Chronic kidney disease or dialysis |
| Chronic liver disease including cirrhosis |
| Chronic neuromuscular disease |
| Chronic obstructive pulmonary disease or emphysema |
| Dementia |
| Diabetes |
| Epilepsy |
| HIV |
| Hypertension |
| Mental health disorders including depression, bipolar, post-traumatic stress disorder, schizophrenia, PTSD |
| Multiple sclerosis |
| Overweight (body mass index ≥25 kg/m$^2$) |

| |
|---|
| Parkinson’s disease |
| Peripheral artery disease |
| Pregnancy |
| Pulmonary fibrosis |
| Pulmonary heart disease |
| Sickle cell disease |
| Thalassemia |

*Conditions presented in Supplementary Table 2 are organized into more discrete groupings. Age, tobacco, and substance use are included separately in propensity score models.

**Additional exclusions applied during matching**

We applied additional exclusions during matching. Due to lack of overlap, persons aged >115 years or who were categorized in the other vaccination category were excluded. Untreated persons who died or were hospitalized on or before their assigned index date were also excluded as were untreated persons who received other outpatient COVID-19 treatments (nirmatrelvir-ritonavir, molnupiravir, sotrovimab, remdesivir) on or prior to the antiviral treatment date.

**Sample size and power calculations**

For comparisons of nirmatrelvir-ritonavir or molnupiravir versus no treatment (trials 1 and 2), given a 1:4 match and assuming a 3% incidence of 30-day hospitalization or death among untreated groups, a sample size of 2,650 persons (530 in treatment, 2,120 in no treatment group) was determined to have 80% power to detect a 2% difference in 30-day hospitalization or death (3% untreated, 1% treated).

For comparisons of nirmatrelvir-ritonavir versus molnupiravir (trial 3), given a 1:1 match and assuming a 3% incidence of 30-day hospitalization or death, a sample size of 826 persons in each group was determined to have 80% power to detect a 2% difference in 30-day hospitalization or death (3% molnupiravir, 1% nirmatrelvir-ritonavir). Calculations are based on a two-way Fisher’s exact test for equality of two independent proportions with a significance level of 0.05.

**Supplemental Table 8. List of conditions evaluated as potential post-COVID incident conditions from day 31 to 180 after the index date***

| Conditions and symptoms attributed to post-COVID illness | Description |
|---|---|
| **Cardiac** | |
| Acute coronary syndrome | Includes unstable angina, NSTEMI, STEMI, myocardial infarction |
| Cardiac dysrhythmias | Includes atrial flutter, atrial fibrillation, tachycardia, atrioventricular block, cardiac arrhythmia |
| Cardiovascular disease | Includes coronary artery disease, peripheral arterial/vascular disease, thoracic/abdominal aneurysm |
| Chest pain | |
| Heart failure and cardiomyopathy | Includes any valvular disease (mitral, aortic, tricuspid, pulmonary), any heart failure, any cardiomyopathy, pulmonary hypertension |
| Hypertension | |
| Myocarditis | |

| | |
|---|---|
| **Pulmonary** | |
| Respiratory symptoms | Includes shortness of breath/dyspnea, any respiratory distress/failure, any bronchitis, hypoxemia, bronchiectasis, any non-COVID pneumonia including influenza, cough, wheezing, sneezing, nasal congestion/sinusitis, sore throat, pharyngitis, laryngitis, tonsillitis. Does not include otitis media. |
| Asthma | |
| COPD and emphysema | |
| Obstructive sleep apnea or obesity hypoventilation | Includes other sleep apnea not specified as central sleep apnea |
| **Renal** | Includes acute kidney injury, chronic kidney disease, dialysis |
| **Hematologic/thromboembolic** | |
| Venous thromboembolism | Includes involvement of any deep veins |
| Pulmonary embolism | |
| **Gastrointestinal** | |
| Abdominal pain | |
| Esophageal disorders | Includes any esophagitis, gastro-esophageal reflux disease, esophageal ulcer |
| Gastrointestinal disorders | Includes any irritable bowel syndrome, inflammatory bowel disease, constipation, diarrhea, nausea, vomiting |
| **Neurologic** | |
| Cerebrovascular disease | Includes stroke; TIA; occlusion of vertebral, basilar, cerebral, cerebellar, or carotid arteries; cerebral aneurysm. Does not include vascular dementia. |
| Dementia | |
| Smell and taste disturbance | |
| Headache | Includes migraine |
| Sleeping disorders | Includes any central or complex sleep apnea, insomnia. Does not include obstructive sleep apnea |
| Other neurologic conditions | Includes peripheral nerve disorders (i.e., neuropathy, Guillan-Barre syndrome), epilepsy, multiple sclerosis, complex regional pain syndrome, Parkinson disease |
| **Cancer** | |
| **Mental Health** | |
| Depression | |
| Other mood disorders | Includes bipolar disorder, schizophrenia, psychosis |
| Anxiety | |
| PTSD | |
| Substance-related disorders | Includes alcohol, cannabis, opioids, stimulants, cocaine |
| **Musculoskeletal** | Includes any myositis, muscle wasting and atrophy, contracture of muscle, myalgias |
| **Endocrine** | |
| Diabetes | Any diabetes excluding gestational diabetes |

| |
|---|
| Disorders of lipid metabolism |
| Obesity |
| **General** |
| Malaise and fatigue |

[*]ICD-10 codes available at: https://github.com/nrajeevan/ICD_codes_post_covid_conditions

### B. SUPPLEMENTAL RESULTS

**Supplemental Table 9. Additional persons excluded from the match-eligible cohort during matching**

| | **Trial #1** | | **Trial #2** | | **Trial #3** | |
|---|---|---|---|---|---|---|
| | **Nirma-trelvir-ritonavir** | **No treatment** | **Molnu-piravir** | **No treatment** | **Molnu-piravir** | **Nirma-trelvir-ritonavir** |
| Match eligible population, N | 1,639 | 103,353 | 922 | 111,233 | 795 | 1,637 |
| **Exclusions, N** | | | | | | |
| Aged >115 years or other vaccination category | <11[*] | 106 | <11[*] | 116 | 0 | 0 |
| Missing Area Deprivation Index | 0 | 169 | 0 | 184 | n/a | n/a |
| Died or were hospitalized on or before assigned index date | n/a | 131 | n/a | 82 | n/a | n/a |
| Received any other outpatient COVID-19 treatment prior to match date | <11[*] | 176 | <11[*] | 162 | n/a | n/a |
| All matches dropped due to hospitalization, death, or treatment prior to assigned index date | <11[*] | n/a | <11[*] | n/a | n/a | n/a |
| No match | 44 | – | 21 | – | 26 | – |

| | | | | | | |
|---|---|---|---|---|---|---|
| Number of unique patients matched, % | 1,587 (97%) | 5,591 | 897 (97%) | 3,213 | 769 (97%) | 534 |

[*]Data from Centers for Medicare & Medicaid Services (CMS) were combined with other data sources and masked in accordance with CMS cell size suppression policy (https://resdac.org/articles/cms-cell-size-suppression-policy)

**Supplemental Table 10. Baseline demographic and clinical characteristics of Veterans who tested positive for SARS-CoV-2 in the VA healthcare system from January 1, 2022 to February 28, 2022, who fulfilled eligibility criteria in three emulated target trials of COVID-19 antiviral effectiveness**

| | Trial 1 | | Trial 2 | | Trial 3 | |
|---|---|---|---|---|---|---|
| **Characteristic[*]** | **Nirmatrelvir-ritonavir** | **No treatment** | **Molnupiravir** | **No treatment** | **Molnupiravir** | **Nirmatrelvir-ritonavir** |
| **Total** | 1,639 | 103,353 | 922 | 111,233 | 795 | 1,637 |
| **Age, years, median [IQR]** | 65.0 (54.0,74.0) | 59.0 (45.0,71.0) | 68.0 (58.0,75.0) | 60.0 (46.0,71.0) | 68.0 (57.0,74.0) | 65.0 (54.0,74.0) |
| **Age group, N (%)** | | | | | | |
| 18-49 | 285(17.4) | 32,751(31.7) | 112(12.1) | 33,642(30.2) | 106(13.3) | 283(17.3) |
| 50-64 | 485(29.6) | 31,794(30.8) | 253(27.4) | 33,965(30.5) | 225(28.3) | 485(29.6) |
| 65-74 | 536(32.7) | 23,726(23.0) | 326(35.4) | 26,505(23.8) | 278(35.0) | 536(32.7) |
| ≥75 | 333(20.3) | 15,082(14.6) | 231(25.1) | 17,121(15.4) | 186(23.4) | 333(20.3) |
| **Male sex, N (%)** | 1,459(89.0) | 88,711(85.8) | 839(91.0) | 96,112(86.4) | 718(90.3) | 1,458(89.1) |
| **Race/Ethnicity, N (%)** | | | | | | |
| Hispanic | 147(9.0) | 10,141(9.8) | 54(5.9) | 10,788(9.7) | 47(5.9) | 147(9.0) |
| White | 1,152(70.3) | 63,058(61.0) | 681(73.9) | 67,777(60.9) | 595(74.8) | 1,150(70.3) |
| Black | 237(14.5) | 21,075(20.4) | 132(14.3) | 22,896(20.6) | 102(12.8) | 237(14.5) |
| Other | 38(2.3) | 3,149(3.0) | 19(2.1) | 3,421(3.1) | 17(2.1) | 38(2.3) |
| Unknown | 65(4.0) | 5,930(5.7) | 36(3.9) | 6,351(5.7) | 34(4.3) | 65(4.0) |
| **Rurality, N (%)[†]** | | | | | | |
| Rural | 302(18.4) | 16,062(15.5) | 204(22.1) | 17,358(15.6) | 179(22.5) | 302(18.4) |
| Urban | 1,337(81.6) | 87,291(84.5) | 718(77.9) | 93,875(84.4) | 616(77.5) | 1,335(81.6) |
| **Region, N (%)[‡]** | | | | | | |
| West | 348(21.3) | 25,899(25.1) | 225(24.4) | 27,793(25.0) | 200(25.2) | 348(21.3) |
| Midwest | 403(24.7) | 18,785(18.2) | 208(22.6) | 20,260(18.3) | 171(21.5) | 403(24.7) |
| Northeast | 328(20.1) | 14,188(13.8) | 98(10.6) | 15,347(13.8) | 81(10.2) | 328(20.1) |
| South | 555(34.0) | 44,287(42.9) | 390(42.3) | 47,609(42.9) | 342(43.1) | 553(33.9) |
| **Area Deprivation Index, median [IQR]** | 59.5 (41.3,76.8) | 56.5 (38.0,73.0) | 61.3 (44.3,77.7) | 56.7 (38.0,73.3) | 63.0 (45.0,78.5) | 59.5 (41.3,76.8) |
| **Month of positive test, N (%)** | | | | | | |
| January | 1,026(62.6) | 86,152(83.4) | 648(70.3) | 92,188(82.9) | 564(70.9) | 1,024(62.6) |
| February | 613(37.4) | 17,201(16.6) | 274(29.7) | 19,045(17.1) | 231(29.1) | 613(37.4) |

| | | | | | | |
|---|---|---|---|---|---|---|
| **Week of positive test, N (%)** | | | | | | |
| January 1-14 | 189 (11.6) | 53,268 (51.6) | 138 (15.0) | 56,563 (50.8) | 121 (15.2) | 188 (11.5) |
| January 15-28 | 742 (45.3) | 31,039 (30.0) | 464 (50.3) | 33,561 (30.1) | 405 (51.0) | 741 (45.3) |
| January 29-February 11 | 474 (28.9) | 13,099 (12.7) | 233 (25.2) | 14,511 (12.9) | 190 (23.9) | 474 (29.0) |
| February 12-28 | 234 (14.3) | 5,947 (5.8) | 87 (9.5) | 6,598 (5.9) | 79 (9.9) | 234 (14.3) |
| **≥1 symptom, N (%)[§]** | | | | | | |
| No | 466(28.4) | 52,089(50.4) | 221(24.0) | 55,522(49.9) | 197(24.8) | 466(28.5) |
| Yes | 1,173(71.6) | 51,264(49.6) | 701(76.0) | 55,711(50.1) | 598(75.2) | 1,171(71.5) |
| **Vaccination status and time since last dose, N (%)[ǁ]** | | | | | | |
| No doses | 474(28.9) | 31,751(30.7) | 207(22.5) | 33,415(30.0) | 190(23.9) | 472(28.8) |
| Partial/>4 months | 64(3.9) | 2,949(2.9) | 25(2.7) | 3,285(3.0) | 24(3.0) | 64(3.9) |
| Partial/0-4 months | 15(0.9) | 1,121(1.1) | 14(1.5) | 1,212(1.1) | 11(1.4) | 15(0.9) |
| Primary/>4 months | 494(30.1) | 36,013(34.8) | 310(33.6) | 38,492(34.6) | 272(34.2) | 494(30.2) |
| Primary/0-4 months | 74(4.5) | 5,147(5.0) | 41(4.4) | 5,612(5.0) | 33(4.2) | 74(4.5) |
| Booster/>4 months | 118(7.2) | 2,853(2.8) | 63(6.8) | 3,298(3.0) | 54(6.8) | 118(7.2) |
| Booster/0-4 months | 399(24.3) | 23,414(22.7) | 261(28.3) | 25,803(23.2) | 210(26.4) | 399(24.4) |
| **NIH Tier, N (%)[ǁ]** | | | | | | |
| 1 | 333(20.3) | 13,049(12.6) | 164(17.8) | 14,651(13.2) | 143(18.0) | 333(20.3) |
| 2 | 288(17.6) | 25,044(24.2) | 119(12.9) | 25,887(23.3) | 113(14.2) | 286(17.5) |
| 3 | 610(37.2) | 27,865(27.0) | 411(44.6) | 31,387(28.2) | 338(42.5) | 610(37.3) |
| 4 | 406(24.8) | 37,028(35.8) | 228(24.7) | 38,933(35.0) | 201(25.3) | 406(24.8) |
| **Smoking, N (%)** | | | | | | |
| Never | 648(39.5) | 40,700(39.4) | 328(35.6) | 43,671(39.3) | 282(35.5) | 647(39.5) |
| Former | 689(42.0) | 40,272(39.0) | 412(44.7) | 43,858(39.4) | 350(44.0) | 689(42.1) |
| Current | 240(14.6) | 17,867(17.3) | 157(17.0) | 18,904(17.0) | 139(17.5) | 239(14.6) |
| Unknown | 62(3.8) | 4,514(4.4) | 25(2.7) | 4,800(4.3) | 24(3.0) | 62(3.8) |
| **Alcohol dependence, N (%)** | 283(17.3) | 23,765(23.0) | 159(17.2) | 25,175(22.6) | 142(17.9) | 281(17.2) |
| **Substance dependence, N (%)** | 56(3.4) | 5,412(5.2) | 39(4.2) | 5,871(5.3) | 31(3.9) | 56(3.4) |
| **Number of underlying conditions** | | | | | | |
| **Median [IQR]** | 4.0 (2.0,6.0) | 3.0 (2.0,5.0) | 4.0 (3.0,14.0) | 3.0 (2.0,5.0) | 4.0 (3.0,13.0) | 4.0 (2.0,6.0) |
| **N (%)** | | | | | | |
| 0-1 | 155(9.5) | 15,758(15.2) | 51(5.5) | 16,098(14.5) | 51(6.4) | 155(9.5) |

| | | | | | | |
|---|---|---|---|---|---|---|
| 2-3 | 634(38.7) | 47,360(45.8) | 283(30.7) | 49,070(44.1) | 262(33.0) | 633(38.7) |
| 4-5 | 371(22.6) | 19,269(18.6) | 216(23.4) | 21,031(18.9) | 199(25.0) | 371(22.7) |
| 5+ | 479(29.2) | 20,966(20.3) | 372(40.3) | 25,034(22.5) | 283(35.6) | 478(29.2) |
| **CAN Score for mortality w/in 1yr at test date (%)¶** | | | | | | |
| 0-30 | 506(30.9) | 46,367(44.9) | 217(23.5) | 47,755(42.9) | 203(25.5) | 504(30.8) |
| 31-55 | 459(28.0) | 25,103(24.3) | 235(25.5) | 26,620(23.9) | 214(26.9) | 459(28.0) |
| 56-75 | 307(18.7) | 15,361(14.9) | 201(21.8) | 16,837(15.1) | 184(23.1) | 307(18.8) |
| 76-90 | 242(14.8) | 10,577(10.2) | 178(19.3) | 12,471(11.2) | 138(17.4) | 242(14.8) |
| 95-96 | 49(3.0) | 1,620(1.6) | 26(2.8) | 2,109(1.9) | 18(2.3) | 49(3.0) |
| 97-98 | 36(2.2) | 1,707(1.7) | 41(4.4) | 2,273(2.0) | 21(2.6) | 36(2.2) |
| 99 | 23(1.4) | 1,095(1.1) | 21(2.3) | 1,524(1.4) | 15(1.9) | 23(1.4) |
| **Underlying condition, N (%)** | | | | | | |
| Obesity (body mass index ≥30 $kg/m^2$) | 850(51.9) | 52,371(50.7) | 498(54.0) | 56,212(50.5) | 420(52.8) | 850(51.9) |
| Chronic kidney disease | 188(11.5) | 9,596(9.3) | 191(20.7) | 13,212(11.9) | 113(14.2) | 188(11.5) |
| Diabetes | 627(38.3) | 28,866(27.9) | 388(42.1) | 32,884(29.6) | 308(38.7) | 627(38.3) |
| Immuno-suppressive medications or cancer therapies‖ | 148(9.0) | 4,582(4.4) | 72(7.8) | 5,387(4.8) | 59(7.4) | 148(9.0) |
| Cancer | 418(25.5) | 17,487(16.9) | 231(25.1) | 19,777(17.8) | 187(23.5) | 418(25.5) |
| Cardiovascular disease | 572(34.9) | 27,334(26.4) | 447(48.5) | 31,834(28.6) | 347(43.6) | 571(34.9) |
| Chronic lung disease | 569(34.7) | 27,831(26.9) | 383(41.5) | 31,013(27.9) | 318(40.0) | 569(34.8) |
| Dementia | 60(3.7) | 3,261(3.2) | 44(4.8) | 3,771(3.4) | 34(4.3) | 60(3.7) |
| Cerebrovascular disease | 81(4.9) | 4,638(4.5) | 86(9.3) | 5,417(4.9) | 64(8.1) | 81(4.9) |
| Chronic liver disease | -- | -- | 5(0.5) | 476(0.4) | -- | -- |
| Mental Health conditions** | 717(43.7) | 52,076(50.4) | 450(48.8) | 55,891(50.2) | 389(48.9) | 715(43.7) |
| Number of healthcare encounters in prior 12 months (mean±SD) | 26.6±24.7 | 23.1±22.5 | 30.9±28.8 | 24.5±24.4 | 27.5±23.1 | 26.6±24.7 |
| Number of healthcare encounters in prior 12 months (median(IQR)) | 20.0 (11.0,35.0) | 17.0 (9.0,29.0) | 23.0 (13.0,38.0) | 17.0 (9.0,31.0) | 22.0 (12.0,35.0) | 20.0 (11.0,35.0) |
| Number of healthcare | | | | | | |

| | | | | | | |
|---|---|---|---|---|---|---|
| encounters in prior 12 months N(%) | | | | | | |
| 0-8 | 289(17.6) | 24,000(23.2) | 124(13.4) | 24,611(22.1) | 119(15.0) | 289(17.7) |
| 9-15 | 323(19.7) | 24,143(23.4) | 163(17.7) | 25,172(22.6) | 155(19.5) | 323(19.7) |
| 16-30 | 533(32.5) | 30,682(29.7) | 306(33.2) | 32,950(29.6) | 271(34.1) | 532(32.5) |
| 30+ | 494(30.1) | 24,528(23.7) | 329(35.7) | 28,500(25.6) | 250(31.4) | 493(30.1) |
| Days from test to treatment N(%) | | | | | | |
| 0-1 | 1349(82.3) | - | 765(83.0) | - | 662(83.3) | 1,347(82.3) |
| 2+ | 290(17.7) | - | 157(17.0) | - | 133(16.7) | 290(17.7) |

*Baseline characteristics represent unweighted persons
†Based on rural-urban commuting area (RUCA) codes
‡Regions are based on Veterans Integrated Service Networks (VISNs). West includes VISNs 19-22; Midwest 10,12,15,23; Northeast 1,2,4,5; South 6-9, 16-17
§Any of 15 pre-specified COVID-19-related symptoms present on the day of positive SARS-CoV-2 test or within the preceding 30 days
‖See Supplemental Appendix. There were 225 persons with other vaccination status and 746 persons with other NIH tier.
¶CAN data missing for 3,206 persons.
**Includes major depressive disorder, bipolar disorder, post-traumatic stress disorder, schizophrenia

**Supplemental Figure 1. Distribution of baseline characteristics and propensity scores in persons who were treated with nirmatrelvir-ritonavir and their comparators receiving no COVID-19 antiviral or monoclonal antibody treatment, before and after matching.**

A. Absolute standardized mean differences and variance ratios of baseline characteristics between nirmatrelvir-ritonavir treatment versus no treatment shown for the raw and matched data.

Prior to matching, the absolute standardized difference in baseline characteristics between the nirmatrelvir-ritonavir treatment versus no treatment groups ranged from 0.003-0.435 with a median of 0.078 (IQR: 0.031-0.147). After matching, the absolute standardized differences ranged from 0.001-0.062 with a median of 0.012 (IQR: 0.005-0.025).

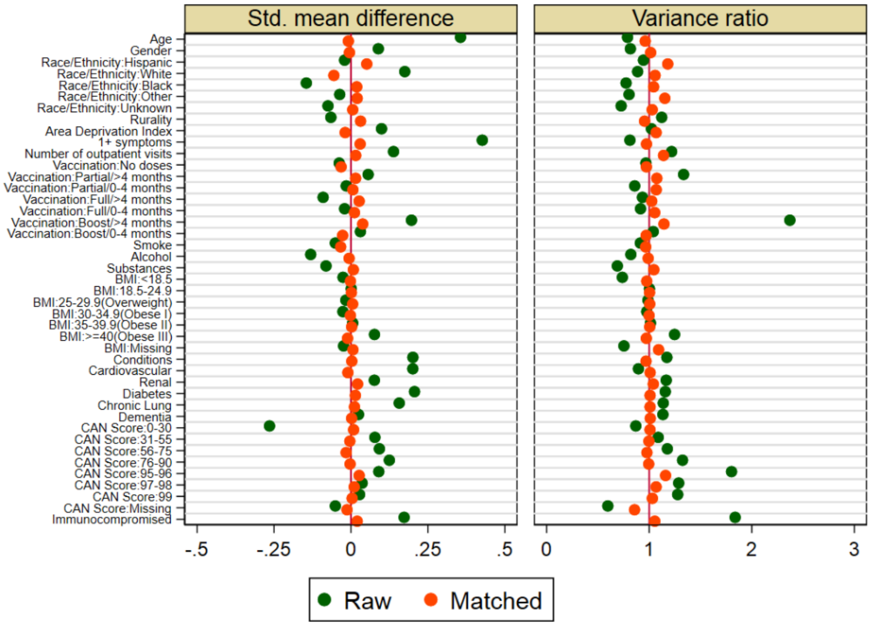

B. Cumulative distribution of propensity score between nirmatrelvir-ritonavir treatment versus no treatment shown for the raw and matched data demonstrate balance after matching.

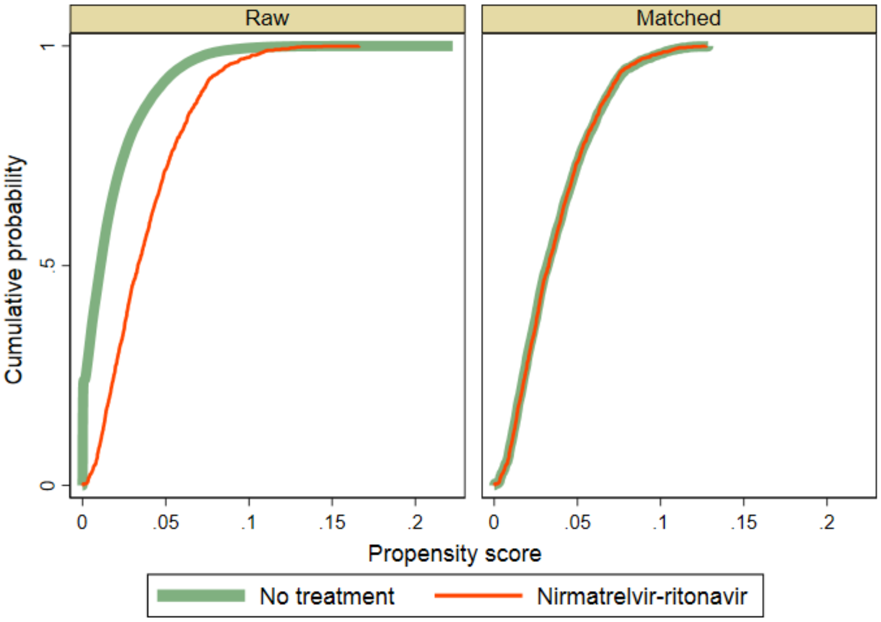

**Supplemental Figure 2. Distribution of baseline characteristics of persons who were treated with molnupiravir and their comparators with no COVID-19 antiviral or monoclonal antibody treatment, before and after matching.**

A. Absolute standardized mean differences and variance ratios of baseline characteristics between molnupiravir treatment versus no treatment shown for the raw and matched data.

Prior to matching, the absolute standardized difference in baseline characteristics between the molnupiravir treatment versus no treatment groups ranged from 0.000-0.526 with a median of 0.118 (IQR: 0.053-0.211). After matching, the absolute standardized differences ranged from 0.001-0.059 with a median of 0.018 (IQR: 0.011-0.032).

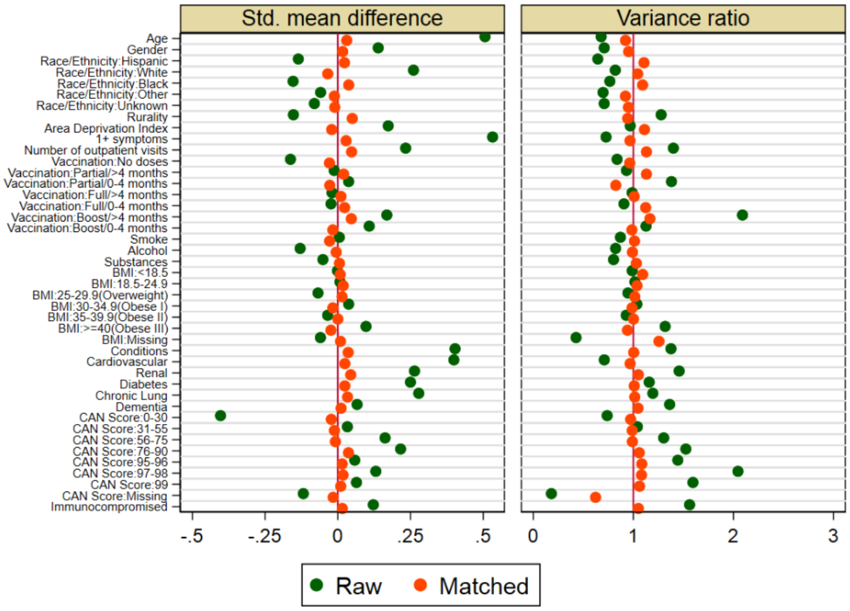

B. Cumulative distribution of propensity score between molnupiravir treatment versus no treatment shown for the raw and matched data demonstrate balance after matching.

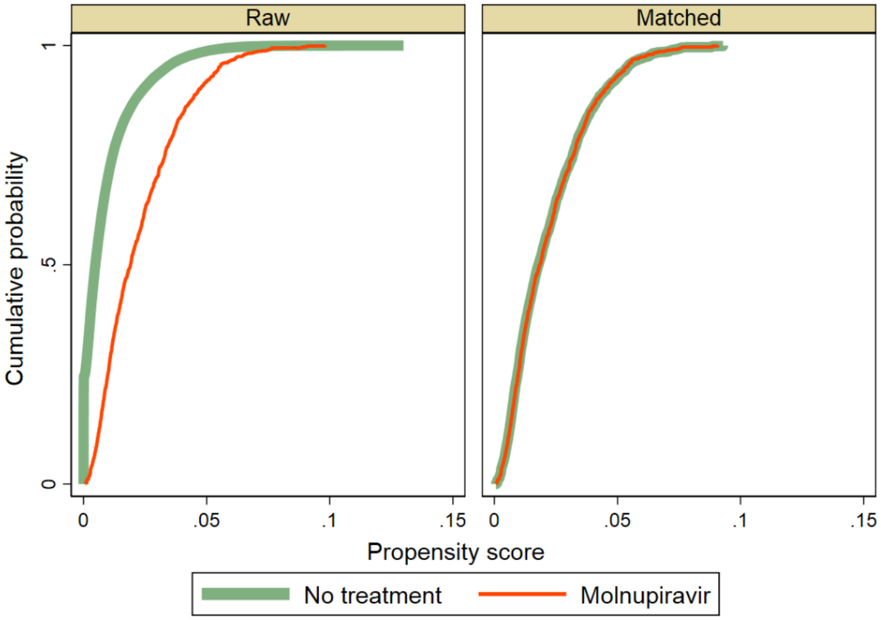

**Supplemental Figure 3. Distribution of baseline characteristics of persons who were treated with molnupiravir and their comparators treated with nirmatrelvir-ritonavir, before and after matching.**

A. Absolute standardized mean differences and variance ratios of baseline characteristics between molnupiravir treatment versus nirmatrelvir-ritonavir treatment shown for the raw and matched data.

Prior to matching, the absolute standardized difference in baseline characteristics between the molnupiravir treatment versus nirmatrelvir-ritonavir treatment groups ranged from 0.001-0.145 with a median of 0.033 (IQR: 0.023-0.079). After matching, the absolute standardized differences ranged from 0.002-0.105 with a median of 0.023 (IQR: 0.011-0.045).

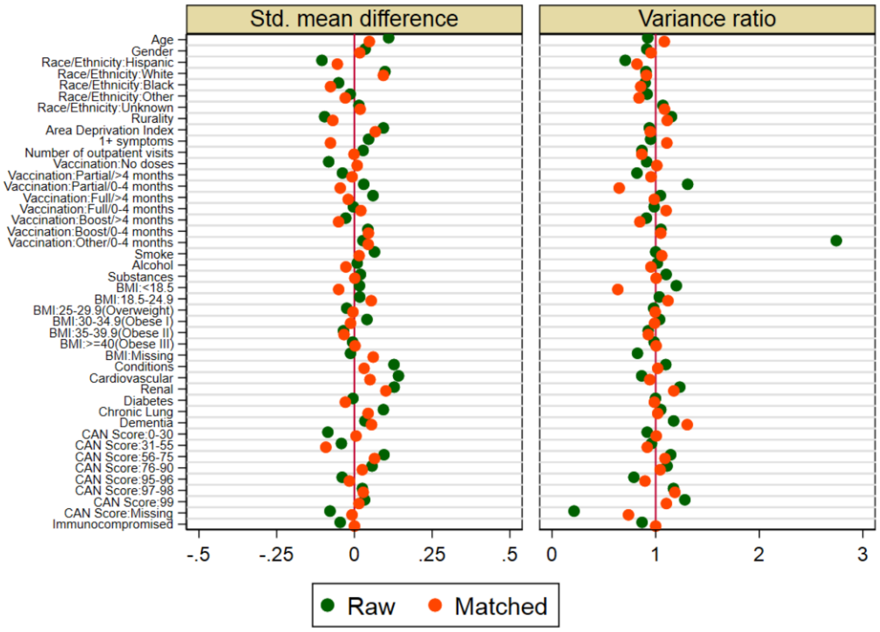

b. Cumulative distribution of propensity score between molnupiravir treatment versus nirmatrelvir-ritonavir treatment shown for the raw and matched data demonstrate balance after matching.

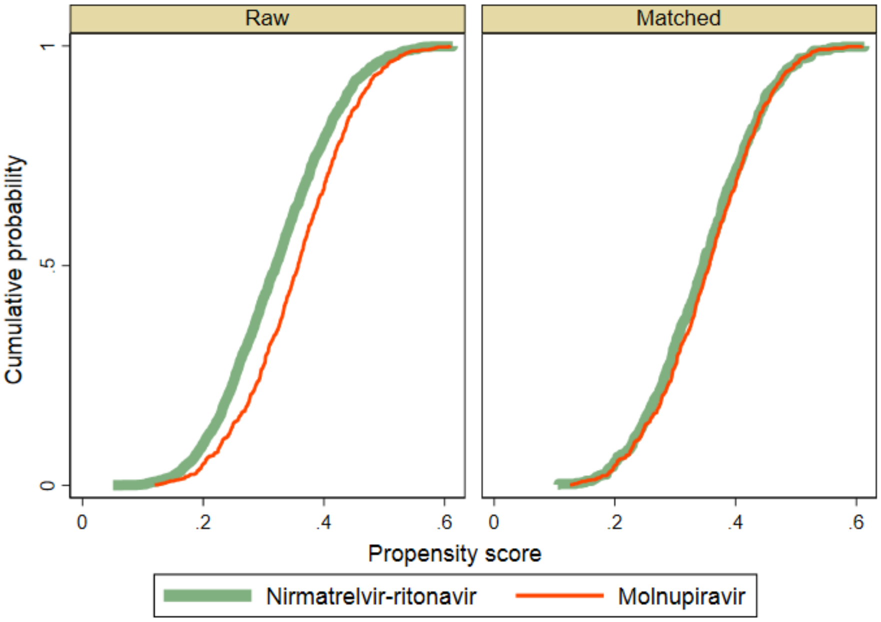

**Supplemental Table 11. Counts of unique and multiply matched Veterans for each of the three target trial emulations**

| | | Trial #1 1:4 | | Trial #2 1:4 | | Trial #3 1:1 | |
|---|---|---|---|---|---|---|---|
| | | Case | Control | Case | Control | Case | Control |
| | | Nirma-trelvir-ritonavir | No treatment | Molnu-piravir | No treatment | Molnu-piravir | Nirma-trelvir-ritonavir |
| **Number of unique patients** | | 1587 | 5591 | 897 | 3213 | 769 | 534 |
| **Number of times a control was matched to a case** | Median | 1 | | 1 | | 1 | |
| | IQR | (1,1) | | (1,1) | | (1,2) | |
| | Range | [1-4] | | [1-3] | | [1-10] | |
| **Number of controls matched to a case** | Median | 4 | | 4 | | 1[*] | |
| | IQR | (4,4) | | (4,4) | | (1,1)[*] | |
| | Range | [1-4] | | [1-4] | | [1-1][*] | |
| **Total number of controls with multiple matches** | N (%) | 544 (9.7%) | | 253 (7.9%) | | 235 (44%) | |

[*]1:1 matching

**Supplemental Table 12. Comparison of matched groups in three emulated target trials of COVID-19 pharmacotherapy with respect to 30-day outcomes of death or hospitalization after index date: subgroup analyses by age, symptoms, and vaccination (primary approach)[†]**

| Subgroup | No. of Events | | 30-day risk, events per 1000 persons (95% CI) | | Risk Difference (95% CI) | Risk Ratio (95% CI) | P-Value* |
|---|---|---|---|---|---|---|---|
| **Trial #1** | **Nirma-trelvir-ritonavir** | **No treatment** | **Nirma-trelvir-ritonavir** | **No treatment** | | | |
| **Age, years** | | | | | | | < 0.001 |
| 18-64 | 15 | 18.3 | 20.19 (12.19-33.25) | 24.99 (19.49-31.98) | -4.80 (-16.85-7.25) | 0.81 ( 0.46-1.42) | < 0.001 |
| ≥65 | 30 | 66.4 | 35.55 (24.94-50.42) | 77.83 (67.83-89.17) | -42.29 (-58.61--25.97) | 0.46 ( 0.31-0.66) | 0.13 |
| **Symptoms in the 30 days prior to positive SARS-CoV-2 test** | | | | | | | < 0.001 |
| 0 | | 27 | 19.52 (10.16-37.18) | 57.43 (47.50-69.27) | -37.90 (-54.79--21.02) | 0.34 ( 0.17-0.67) | 0.41 |
| ≥1 | 36 | 57.8 | 31.97 (23.14-44.03) | 51.71 (44.31-60.26) | -19.74 (-32.71--6.76) | 0.62 ( 0.43-0.88) | < 0.001 |
| **COVID-19 vaccination status** | | | | | | | < 0.001 |
| Unvaccinated | 20 | 34 | 43.38 (28.11-66.39) | 70.96 (59.03-85.07) | -27.57 (-50.20--4.95) | 0.61 ( 0.38-0.97) | < 0.001 |
| Primary or booster vaccination | 25 | 50.8 | 22.20 (15.04-32.67) | 45.81 (39.01-53.73) | -23.61 (-34.86--12.35) | 0.48 ( 0.32-0.73) | < 0.001 |
| **Trial #2** | **Molnu-piravir** | **No treatment** | **Molnu-piravir** | **No treatment** | | | |
| **Age, years** | | | | | | | < 0.001 |
| 18-64 | 10 | 7.6 | 28.25 (15.22-51.84) | 21.14 (14.63-30.46) | 7.11 (-12.17-26.38) | 1.34 ( 0.64-2.77) | < 0.001 |
| ≥65 | 30 | 44.3 | 55.25 (38.85-78.01) | 82.35 (70.07-96.56) | -27.10 (-50.63--3.58) | 0.67 ( 0.46-0.99) | < 0.001 |
| **Symptoms in the 30 days prior to positive SARS-CoV-2 test** | | | | | | | 0.17 |
| 0 | 6 | 10.8 | 27.40 (12.28-59.99) | 48.82 (36.12-65.68) | -21.42 (-47.44-4.60) | 0.56 ( 0.24-1.30) | 0.22 |

| | | | | | | | |
|---|---|---|---|---|---|---|---|
| ≥1 | 34 | 41.1 | 50.15 (36.02-69.42) | 60.86 (51.31-72.05) | -10.71 (-30.47-9.05) | 0.82 ( 0.57-1.20) | 0.37 |
| **COVID-19 vaccination status** | | | | | | | < 0.01 |
| Unvaccinated | 14 | 17.3 | 69.65 (41.50-114.63) | 82.05 (63.81-104.92) | -12.40 (-53.33-28.53) | 0.85 ( 0.48-1.49) | < 0.01 |
| Primary or booster vaccination | 26 | 34.6 | 37.36 (25.53-54.35) | 50.43 (41.92-60.56) | -13.08 (-29.99-3.84) | 0.74 ( 0.49-1.13) | < 0.01 |
| **Trial #3** | **Nirma-trelvir-ritonavir** | **Molnu-piravir** | **Nirma-trelvir-ritonavir** | **Molnu-piravir** | | | |
| **Age, years** | | | | | | | 0.12 |
| 18-64 | 5 | 8 | 15.53 ( 5.45-43.44) | 25.32 (12.66-49.98) | -9.79 (-33.60-14.03) | 0.61 ( 0.18-2.14) | 0.11 |
| ≥65 | 14 | 22 | 31.32 (16.71-57.94) | 48.57 (32.14-72.76) | -17.25 (-44.75-10.26) | 0.64 ( 0.31-1.35) | 0.52 |
| **COVID-19 vaccination status** | | | | | | | 0.03 |
| Unvaccinated | 6 | 13 | 32.61 (12.76-80.82) | 71.43 (41.71-119.68) | -38.82 (-87.70-10.06) | 0.46 ( 0.16-1.34) | 0.01 |
| Primary or booster vaccination | 13 | 17 | 22.22 (11.49-42.54) | 28.96 (18.05-46.15) | -6.74 (-26.18-12.71) | 0.77 ( 0.35-1.69) | 0.02 |

*Calculated using Wald test for interaction using the risk ratio models.
†Analysis is conducted according to the appropriate subgroup of case-participants. Matched control-participants may belong to any subgroup (see Supplemental Table 13 for secondary approach).

**Supplemental Table 13. Comparison of matched groups in three emulated target trials of COVID-19 pharmacotherapy with respect to 30-day outcomes of death or hospitalization after index date: subgroup analyses by age, symptoms, and vaccination (secondary approach)†**

| Subgroup | No. of Events | | 30-day risk, events per 1000 persons (95% CI) | | Risk Difference (95% CI) | Risk Ratio (95% CI) | P-value* |
|---|---|---|---|---|---|---|---|
| **Trial #1** | **Nirma-trelvir-ritonavir** | **No treatment** | **Nirma-trelvir-ritonavir** | **No treatment** | | | |
| **Age, years** | | | | | | | < 0.001 |
| 18-64 | 12 | 17 | 16.97 ( 9.65-29.69) | 24.05 (17.97-32.10) | -7.07 (-19.11-4.97) | 0.71 ( 0.37-1.34) | < 0.001 |

| | | | | | | | |
|---|---|---|---|---|---|---|---|
| ≥65 | 30 | 65.5 | 35.55 (24.94-50.42) | 77.61 (66.48-90.41) | -42.06 (-59.29--24.84) | 0.46 ( 0.31-0.67) | 0.06 |
| **Symptoms in the 30 days prior to positive SARS-CoV-2 test** | | | | | | | < 0.001 |
| 0 | 8 | 24.8 | 18.60 ( 9.31-36.85) | 57.75 (45.72-72.70) | -39.15 (-58.11--20.18) | 0.32 ( 0.15-0.67) | 0.68 |
| ≥1 | 35 | 60 | 31.62 (22.77-43.74) | 54.20 (45.49-64.46) | -22.58 (-36.22--8.95) | 0.58 ( 0.41-0.84) | < 0.001 |
| **COVID-19 vaccination status** | | | | | | | < 0.001 |
| Unvaccinated | 20 | 34.4 | 43.48 (28.17-66.53) | 74.82 (59.84-93.17) | -31.34 (-56.01--6.67) | 0.58 ( 0.36-0.94) | < 0.001 |
| Primary or booster vaccination | 24 | 49.7 | 22.20 (14.91-32.93) | 45.95 (38.42-54.86) | -23.74 (-35.74--11.75) | 0.48 ( 0.31-0.74) | < 0.001 |
| **Trial #2** | **Molnu-piravir** | **No treatment** | **Molnu-piravir** | **No treatment** | | | |
| **Age, years** | | | | | | | < 0.001 |
| 18-64 | 10 | 7.6 | 28.99 (15.62-53.18) | 21.98 (13.57-35.42) | 7.00 (-13.93-27.94) | 1.32 ( 0.60-2.90) | < 0.001 |
| ≥65 | 30 | 44.6 | 55.35 (38.92-78.15) | 82.26 (69.85-96.64) | -26.91 (-50.58--3.24) | 0.67 ( 0.46-0.99) | < 0.01 |
| **Symptoms in the 30 days prior to positive SARS-CoV-2 test** | | | | | | | 0.09 |
| 0 | 4 | 11 | 19.42 ( 7.25-50.98) | 53.40 (37.00-76.49) | -33.98 (-60.88--7.08) | 0.36 ( 0.13-1.02) | 0.42 |
| ≥1 | 34 | 42.5 | 50.60 (36.34-70.04) | 63.24 (53.25-74.97) | -12.65 (-32.71-7.42) | 0.80 ( 0.55-1.16) | 0.15 |
| **COVID-19 vaccination status** | | | | | | | 0.02 |
| Unvaccinated | 13 | 15.7 | 65.99 (38.52-110.80) | 79.53 (56.52-110.79) | -13.54 (-59.04-31.97) | 0.83 ( 0.44-1.58) | 0.03 |

| | | | | | | | |
|---|---|---|---|---|---|---|---|
| Primary or booster vaccination | 24 | 33.3 | 35.61 (23.95-52.63) | 49.46 (40.32-60.53) | -13.85 (-30.69-2.99) | 0.72 ( 0.47-1.11) | 0.01 |
| **Trial #3** | **Nirma-trelvir-ritonavir** | **Molnu-piravir** | **Nirma-trelvir-ritonavir** | **Molnu-piravir** | | | |
| **Age, years** | | | | | | | 0.17 |
| 18-64 | 4 | 8 | 13.25 ( 3.94-43.57) | 26.49 (13.25-52.26) | -13.25 (-37.53-11.04) | 0.50 ( 0.12-2.00) | 0.2 |
| ≥65 | 14 | 20 | 32.33 (17.26-59.77) | 46.19 (29.94-70.62) | -13.86 (-41.76-14.05) | 0.70 ( 0.33-1.48) | 0.14 |
| **COVID-19 vaccination status** | | | | | | | 0.06 |
| Unvaccinated | 6 | 9 | 41.10 (16.10-100.94) | 61.64 (32.13-115.04) | -20.55 (-75.97-34.88) | 0.67 ( 0.21-2.07) | 0.03 |
| Primary or booster vaccination | 10 | 14 | 18.21 ( 8.72-37.67) | 25.50 (15.13-42.66) | -7.29 (-25.51-10.94) | 0.71 ( 0.30-1.71) | 0.05 |

*Calculated using Wald test for interaction using the risk ratio models.
†Analysis is conducted according to the appropriate subgroup of case-participants. Matched control participants not belonging to the subgroup of interest are dropped, and remaining controls are reweighted (see Supplemental Table 12 for primary approach).

**Supplemental Table 14. Comparison of matched groups in three emulated target trials of COVID-19 pharmacotherapy among Veterans who tested positive for SARS-CoV-2 from January 1, 2022 to February 28, 2022 with respect to cumulative 0-180 day incidence of hospitalization, death, and post-COVID conditions**

| | **Trial 1 Nirmatrelvir-ritonavir vs. No Treatment 31-180 day incidence** per 1000 persons | | **HR or SHR* (95% CI)** | **Trial 2 Molnupiravir vs. No Treatment 31-180 day incidence** per 1000 persons | | **HR or SHR* (95% CI)** | **Trial 3 Nirmatrelvir-ritonavir vs. Molnupiravir 31-180 day incidence** per 1000 persons | | **HR or SHR* (95% CI)** |
|---|---|---|---|---|---|---|---|---|---|
| **31-180 day outcome** | Nirma-trelvir-ritonavir N=1587 | No treat-ment N=1587 | | Molnupiravir N=897 | No treat-ment N=897 | | Nirma-trelvir-ritonavir N=769 | Molnu-piravir N=769 | |
| Acute or long-term care admission or death | 118.8 | 144.0 | 0.82 (0.69-0.98) | 190.8 | 163.8 | 1.16 (0.96-1.41) | 140.4 | 176.4 | 0.80 (0.57-1.11) |

| | | | | | | | | | |
|---|---|---|---|---|---|---|---|---|---|
| Acute or long-term care admission | 113.4 | 122.4 | 0.93 (0.745 -1.16) | 180.0 | 138.6 | 1.30 (1.01-1.68) | 135.0 | 163.8 | 0.83 (0.60-1.15) |
| Death | 12.6 | 30.6 | 0.42 (0.26-0.67) | 27.0 | 32.4 | 0.82 (0.52-1.29) | 12.6 | 27.0 | 0.50 (0.22-1.10) |

*Sub-hazard ratios (SHR), derived from proportional hazards regression accounting for the competing risk of death, are presented for acute or long-term care admissions and post-COVID outcomes. Hazard ratios are presented for death and acute or long-term care admission or death.

**Supplemental Figure 4. Kaplan-Meier curves comparing Veterans treated with nirmatrelvir-ritonavir versus their matched untreated counterparts (trial 1), molnupiravir versus their matched untreated counterparts (trial 2), and nirmatrelvir-ritonavir versus molnupiravir (trial 3) showing cumulative incidence (%) with 95% confidence intervals of any hospitalization or all-cause death, any hospitalization, and all-cause death 0-180 days after the index date among match groups.**

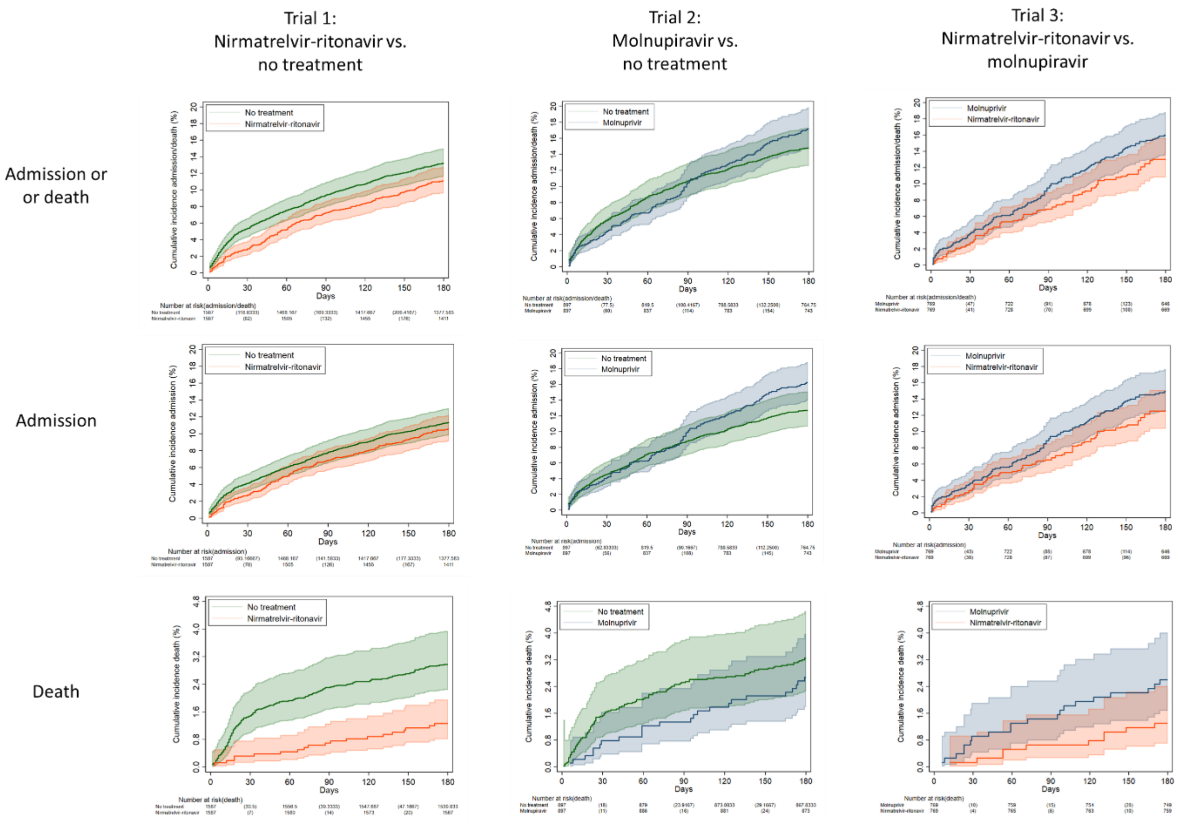
